# Structural brain correlates of depression and anxiety in middle-aged and older adults: A systematic review

**DOI:** 10.1101/2024.08.20.24312289

**Authors:** Sasha Johns, Asri Maharani, Caroline Lea-Carnall, Nick Shryane

## Abstract

Depression and anxiety are associated with structural brain changes, however, research in middle-aged and older adults is inconclusive regarding the regions implicated and the magnitude or direction of changes. We hypothesised that the impact on brain structure may be greater in this age group. We conducted a systematic review on the associations between brain structure with depression and anxiety separately to determine: i) if there are brain regions consistently implicated in depression and/or anxiety, ii) if these are the same for depression and anxiety or distinct, and iii) the extent to which research has dealt with both disorders together. Brain regions consistently associated with depression include the hippocampus, orbitofrontal cortex, prefrontal cortex, insula and temporal lobe. Relationships were mostly negative, with diminished brain structure being associated with increased depression. We were unable to identify regions consistently implicated in anxiety due to a lack of studies investigating its links with brain structure. This review has highlighted the urgent need for research into brain structure and anxiety in middle-aged and older adults.

## INTRODUCTION

### The impact of poor mental health

Poor mental health is a significant worldwide issue that is growing in its impact. The Global Burden of Diseases, Injuries, and Risk Factors Study (GBD) estimated the disability-adjusted life years (DALYs) due to mental disorders increased from 80.8 million (3.1% of total DALYs) in 1990 to 125.3 million (4.9% of total DALYs) in 2019 (GBD 2019 Mental Disorders Collaborators, 2022). These numbers, however, may be underestimated as Arias et al. (2022) predicted that 418 million DALYs (16% of total DALYs) could be attributable to mental disorders after taking into account additional diseases, including neurological disorders. They estimated that the economic impact of this global burden is approximately USD 5 trillion (Arias et al., 2022). The Adult Psychiatric Morbidity Survey, 2014, found that one in six people aged 16 and older in England (17%) had symptoms of a common mental disorder (CMD) in the week prior to being surveyed (McManus et al., 2016). “Common mental disorders” refers to conditions such as depression, anxiety, panic disorder, phobias and obsessive-compulsive disorder (Baker & Kirk-Wade, 2024).Generalised anxiety disorder was the highest reported CMD, at 5.9%, followed by depressive episodes (3.3%) (McManus et al., 2016).

The two most common mental disorders across sex and year were depressive and anxiety disorders, which made up 37.3% and 22.9% of DALYs due to mental disorders in 2019, respectively (GBD 2019 Mental Disorders Collaborators, 2022). The global prevalence of mental health problems has purportedly increased due to the COVID-19 pandemic, particularly among health care workers, non-infectious chronic disease patients, COVID-19 patients, and people in quarantine (Wu et al., 2021). A systematic review and meta-analysis found overall pooled prevalence of depression and anxiety was 31.4% and 31.9% during the COVID-19 pandemic, respectively, which is much higher than the rates reported in previous studies (Wu et al., 2021). Similarly, a study in the UK reported that the prevalence of common mental health problems (assessed using the PHQ-9 and GAD-7) during the COVID-19 lockdown was 52%, compared to around 17% before the pandemic (Pieh et al, 2021).

Individuals with poor mental health also have a much higher prevalence of physical illness. Mental health issues contribute to higher incidence of, and increased mortality from, many health conditions such as heart disease, stroke, diabetes, infections and respiratory disease (Friedli, 2009) in addition to other factors such as obesity, alcohol misuse and smoking (Royal College of Psychiatrists, RSPSYCH, 2010). This relationship also works the other way, with poor physical health increasing the risk of mental illness (RSPSYCH, 2010). They are also likely to receive a reduced standard of care (De Hert et al., 2011).

In addition to the impact on the individual, poor mental health has significant societal and economic implications, partly due to the loss of economic productivity (Kirkwood et al., 2010). The World Health Organization (WHO) produced a factsheet outlining the undefined societal and economic burden that poor mental health imposes. These include but are not limited to: lost production from premature deaths from suicide; lost production from individuals with mental illnesses not being able to work; lost productivity due to family members caring for people who are mentally ill; unemployment, alienation and crime in young people whose mental health issues have caused them to be unable to access education effectively; and emotional burden and reduced quality of life for family members (WHO, 2001).

Depression and anxiety commonly co-occur, they have one of the highest rates of comorbidity of all psychiatric diagnoses (Kessler et al., 2005). It is estimated that up to 40-50% of individuals with major depressive disorder (MDD) also suffer from anxiety (Xia et al., 2018). In addition to the co-occurrence of the disorders, anxious mood has been reported to correspond to depressed mood temporally on a daily basis (Starr & Davila, 2012). When depression is comorbid with anxiety, it is associated with worse health outcomes than having either depression or anxiety on their own (Mittal et al., 2006).

### Depression and Anxiety in Older Adults

Within the UK and globally, we have an ageing population. There is growth in both the size and proportion of older people in each country in the world (WHO, 2022). By 2030, it is estimated that 1 in 6 people in the world will be aged 60 or over. It is expected that the number of people aged 80 or over will triple between 2020 and 2050, reaching 426 million (WHO, 2022). The UK 2021 census reported that the number of people aged 65 or over has increased since the 2011 census. There are now over 11 million people in this age group, which is 18.6% of the total population. In 2011, 16.4% of the total population were 65 or over (Office for National Statistics, 2022). Depression in older adults is a public health concern that is associated with increased suffering, healthcare costs, suicide and mortality related to other causes (Vasiliadis et al., 2013; Aziz & Steffens, 2013; Jiang et al., 2020). MDD is less common in late-life, however depression is associated with a more chronic course in older compared with middle-aged adults. This is likely moderated by medical comorbidity (Haigh, et al., 2018). There is also evidence that depression worsens with age, with depression at older ages being associated with cognitive deficits and poor treatment response (Özel et al., 2022).

Although there is a significant amount of research into depression in older adults, there is an urgent need for more research into anxiety disorders in this population (Byrne & Pachana, 2010). Anxiety is more common than depression in later life, but is studied less (Blay & Marinho, 2012). It has also been suggested that the DSM-IV criteria for an anxiety disorder does not take into consideration the unique characteristics of anxiety in older age and do not allow for discrimination of anxious from non-anxious older adults (Kogan et al., 2000). The burden and prevalence of depression have steadily increased as a result of population growth and ageing (Whiteford et al., 2013; World Health Organization; 2017). Given that this current course is expected to continue, and the known detrimental impact of depression and anxiety on both the individual and society, improving understanding of the mechanisms involved in these disorders is of vital importance.

### Depression and Anxiety and Brain Structure

Research has shown evidence of brain changes, both structural and functional, in individuals with depression and anxiety (e.g. Laird et al., 2019; Maggioni et al., 2019; Peng et al., 2019; Favaro et al., 2015; Shi et al., 2020; Paul et al., 2019; Peng et al., 2018). Depression and anxiety are currently believed to arise from dysregulations of neuroendocrine, neurotransmitter and neuroanatomical systems relating to gene-environment interactions (aan het Rot et al., 2009; Martin et al., 2009; Nemeroff & Vale, 2005; Palazidou, 2012). Observed alterations in brain structure, function or neurotransmitter signalling may be the result of a combination of environmental experiences and genetic predisposition. These alterations then pose an increased risk of psychopathology (Martin et al., 2009). Late-life depression (LLD) is associated with the disruption of brain networks involved in emotional regulation and cognitive processing due to structural connectivity impairments. LLD has been shown to be associated with neuroimaging metrics such as white matter hyperintensity burden, white matter integrity and cortical volume and thickness (Kim & Han, 2021), which are markers of structural brain integrity. Anxiety in later life has been associated with disrupted brain structure and function, hypothalamic-pituitary-adrenal (HPA) axis dysregulation and variation in the serotonin transporter gene (5-HTTLPR) (Blay & Marinho, 2012).

There is evidence that neuroimaging can be utilised for the diagnosis and treatment of depression (Wise et al., 2014). Magnetic Resonance Imaging (MRI) specifically may also assist in the prediction of an individual’s treatment response and illness trajectory in MDD (Alves et al., 2014). Recent developments in neuroimaging techniques have also shown utility in identifying the neurobiological basis of anxiety and contributed to its diagnosis and treatment (Paulus, 2008). It has also been suggested that depression is more complex in older adults, and as treatment is often ineffective, it would be beneficial to use neuroimaging to identify organic brain changes and adjust treatment plans accordingly (Wu et al., 2023). It has also been suggested that improving outcomes in late-life mood and anxiety disorders is a particular challenge, and that neuroimaging techniques may be able to be used to offer “personalised pharmacotherapy” or identifying dysfunctional regions or networks to be the target of direct interventions such as transcranial magnetic stimulation (Ly & Andreescu, 2018).

This review focuses on structural correlates as brain structure may be more fixed and less open to influence from task or emotional state than other measures such as brain function. Brain structure is subject to change, evidenced by the fact that brain structure can change following therapeutic interventions (Malhotra & Sahoo, 2017; Collerton, 2013; Hölzel et al., 2011), exercise (Bugg & Head, 2011; Erickson et al., 2011), and medication (Guo et al., 2015; Younger et al., 2011), but on a slower time frame. Due to the lower rate of change when compared with brain function, any differences in brain structure that are found to relate to depression and anxiety are more likely to be true neural correlates of trait anxiety or depression, rather than state anxiety or low mood on the day of scanning. One could argue that brain structure would be a more constant, measurable analogue of depression and anxiety than brain function and therefore could provide useful information for improving the burden of anxiety and depression discussed above in terms of diagnosis, prognosis and treatment.

Although there is growing evidence of structural changes in the brains of individuals with depression and anxiety (e.g. Zhang et al., 2018; Besteher et al., 2017; Chen et al., 2020; Bas-Hoogendam et al., 2017), there is no current consensus for direction or magnitude of these effects. For example, some studies report diminished structural integrity in depression and anxiety such as reduced grey matter volume (Kandilarova et al., 2019; Ma et al., 2019) or cortical thinning (Bos et al., 2018; Gaspersz et al., 2018) and others find both increased and decreased grey matter volume and cortical thickness, depending on the region (Ancelin et al., 2019; Liao et al., 2011; Gold et al., 2017). However, much of this research discussed focuses on younger adults or adolescents (Besteher et al., 2017; Bas-Hoogendam et al., 2017; Bos et al., 2018; Liao et al., 2011; Gold et al., 2017), and given that there is evidence that depression and anxiety at older ages may be more complex with worse treatment outcomes, the findings may not apply to middle-aged and older adults in the same way. As described previously, depression and anxiety are often comorbid, with a cooccurrence rate as high as 50%. Given this fact, it is essential that research accounts for, or excludes anxiety disorders when examining structural brain changes in depression, and vice versa. Indeed, anxiety has been shown to modulate differences in brain structure seen in depression (Espinoza Oyarce et al., 2020).

### The Aim of the Review

In order to examine the existing literature on structural areas implicated in depression and anxiety in middle-aged and older adults, we conducted a systematic review. We hypothesise that there may be compounding effects of depression and anxiety with ageing, and the impact on brain structure may be greater in middle-aged and older adults. For example, it has been reported that ageing- and disease-related processes can damage the integrity of frontostriatal pathways, the amygdala and the hippocampus, leading to increased vulnerability to depression (Alexopolous, 2005). Also, accelerated brain ageing has been associated with MDD and anxiety, particularly somatic depressive symptoms (Han et al., 2021). In this age group, there are additional variables that need to be considered such as age of onset, number of episodes, duration of symptoms, medication use (Szymkowicz et al., 2023; Korten et al., 2012; Linnebur er al., 2014). It has been suggested that late-onset depression is a “different disorder” and may need to be studied separately from MDD (Zhukovsky et al., 2021).

We aimed to establish if there were any neural regions consistently implicated in depression and anxiety. We also aimed to identify whether common or distinct neural areas are implicated in depression and anxiety. We are also interested in whether studies into depression control for anxiety and vice versa, given their intercorrelation (Eysenck & Fajkowska, 2018; Piccinelli, 1998; Unick et al., 2009). The overall aim of the review is to summarise the current literature to guide future research.

### Research questions

Three research questions were formulated from the objectives of the systematic review.

RQ1- Are there any brain regions that are consistently implicated in depression and anxiety in middle aged and older adults?

RQ2- Are these brain regions common to both depression and anxiety, or are they distinct?

RQ3- To what extent has research dealt with both depression and anxiety together?

## METHODS

### Search strategy and selection criteria

We conducted a search of the APA PsycInfo, Embase & OvidMEDLINE(R) databases on the 7^th^ August 2023 including data from inception to 7^th^ August 2023. In line with objectives, the titles and/or abstracts were required to include the search “anxi*” OR “depress*”, and one from each of the following categories: 1) “imaging” OR “MRI” OR “DTI”, 2) “neuro*” OR “brain” and 3) “middle-age*” OR “older adult*”. Search results were limited to the English language. We designed the selection strategy to produce a report that was most representative of research conducted into structural neuroimaging, in depression and anxiety, in middle-aged and older adult participants who were otherwise healthy. The target population included any adults of the appropriate age who had structural brain imaging and completed depression and/or anxiety questionnaires. Individuals did not have to have a diagnosis of or meet diagnostic criteria for depression or anxiety as we intended to include sub-clinical levels of depression and anxiety.

We chose structural MRI and Diffusion Tensor Imaging (DTI) as the review focused on brain structure. Structural MRI is a brain scanning technique that is used across various clinical and research purposes due to its excellent anatomical detail and strong grey and white matter contrast. It allows measurement of anatomical location, cortical thickness, regional volumes, vascular lesions, and morphological alterations (DiProspero et al., 2022). DTI is a technology used for imaging the white matter of the brain by measuring the diffusion of water molecules. It measures diffusion anisotropy which refers to how tissue varies with direction, thus allowing underlying tissue orientation to be measured (O’Donnell & Westin, 2011). It can be used to identify microstructural changes or neuropathology. There are a number of resulting DTI measures such as mean diffusivity (MD), and fractional anisotropy (FA, Alexander et al., 2007).

We used the Ovid deduplication tool to remove duplicate papers. Any remaining duplicated papers were removed manually.

We designed the review in accordance with the Preferred Reporting Items for Systematic Review and Meta-Analysis guidelines (PRISMA). PRISMA checklist is available in Appendix-Table A. Rayyan software for systematic reviews was used. The 730 articles were imported and the deduplication tool within the software was used. 41 duplicates were removed.

### Eligibility criteria, screening and article selection

We used the American Psychological Association (APA, n.d.) definition of middle-aged as between 36-65 years old. Papers with any participants below this age were excluded. Only English language articles and those with a full manuscript available were included.

### Quality assessment

In order to ensure the quality of each included study, at each stage the first reviewer checked each paper against the inclusion and exclusion criteria with the second reviewer assessing 10% of papers. The second reviewer at each stage was one of the other authors.

The first reviewer examined each paper and coded them for variables such as number of participants, population, mean age, what type of structural imaging was used, the depression or anxiety scales used, whether analysis was whole brain or region of interest (ROI), what measure of brain structure was used and what brain findings were reported.

## RESULTS

### Study selection

The initial search, after deduplication, produced 689 papers. We screened these papers by title, then abstract, then full text (see Figure 1 for a flow diagram of the search and selection process). We recorded reasons for exclusion at the full-screen level. As the review was concerned with brain structure only, we excluded all neuroimaging modalities at the full screen apart from structural MRI and DTI. We focused only on studies considering depression and/or anxiety, rather than any other disorders that may have an effect on brain structure. This process resulted in a final 62 papers that met the criteria (see Table 1, N.B. these papers are included in the reference list and indicated with a *).

**Figure 1:**
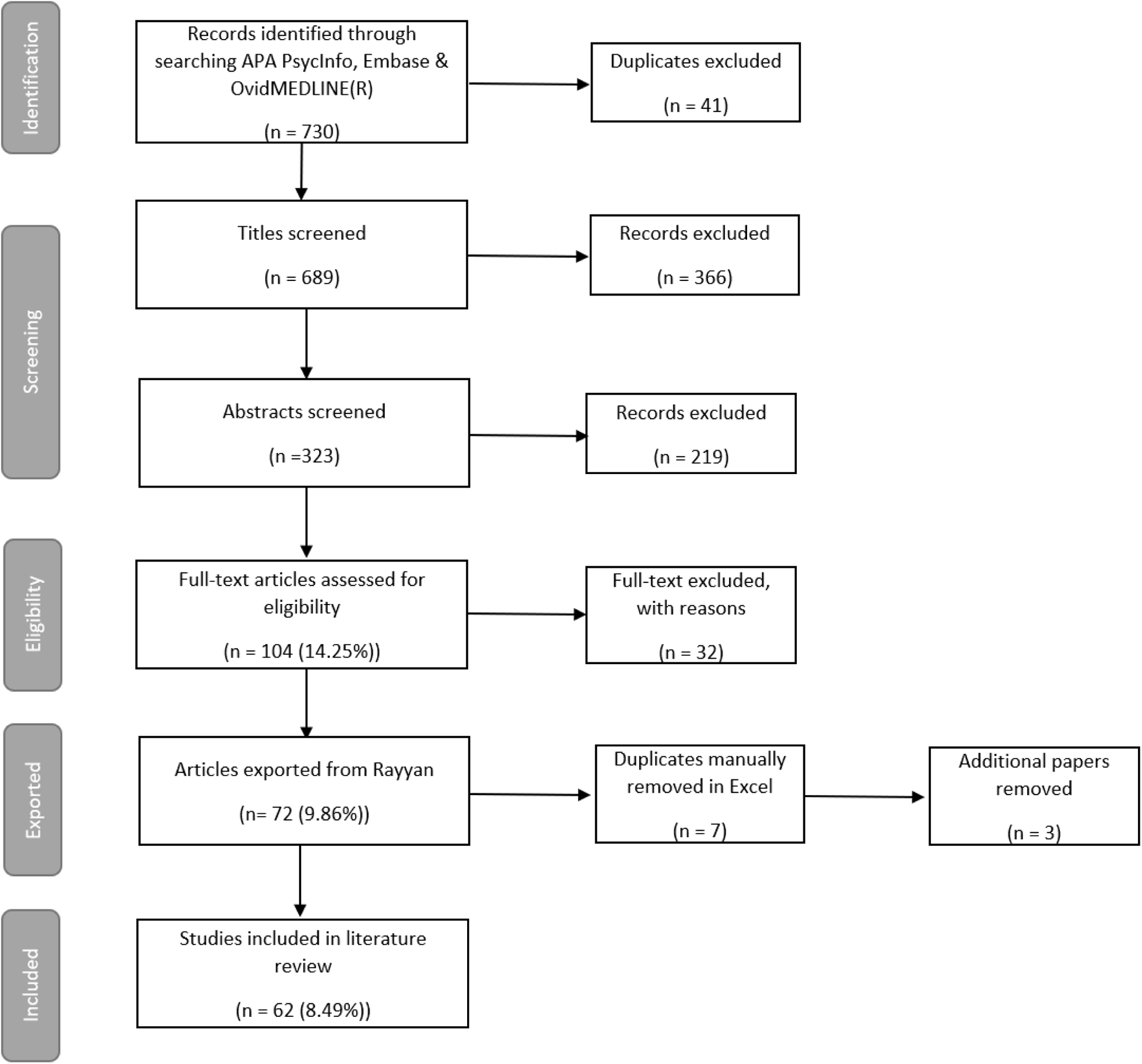
Flow diagram illustrating the literature search and selection process used.

**Table 1:** details of the 62 papers kept in the final review.

| Author<br>(Year) | Sample<br>N=<br>Mean Age<br>Clinical<br>population<br>Y/N | Measured<br>Depression<br>Y/N<br>(*Depression*<br>*Measure*)<br>Measured<br>Anxiety<br>Y/N<br>(*Anxiety*<br>*Measure*) | Dep brain<br>analysis<br>Anx brain<br>analysis | MRI/<br>DTI | Brain<br>Measure | Whole<br>Brain<br>/ROI | Controlled<br>for<br>anx/dep | Medication | No. of<br>episodes | Age<br>at<br>Onset | Symptom<br>duration | All | Sig.<br>brain<br>findings<br>Y/N |
| --- | --- | --- | --- | --- | --- | --- | --- | --- | --- | --- | --- | --- | --- |
| Schuurmans<br>et al. (2023) | 1673<br>47.6<br>N | Y (*BSI*<br>*depressive*<br>*subscale*)<br>N | Y<br>N | MRI | Grey matter volume,<br>white matter volume,<br>white matter lesions,<br>subcortical volume,<br>cortical thickness and<br>cortical surface area | Whole<br>brain | N | N | N | N | Y | N | Y |
| Wu et al.<br>(2023) | 37<br>59<br>Y | Y (*HAMD*)<br>N | Y<br>N | MRI | Grey matter volume | Whole<br>brain | N | Y | N | Y | N | N | Y |
| Espinosa et<br>al. (2022) | 118<br>63.55<br>Y | Y (*GDS-15*<br>*+ DSM-IV*)<br>N | Y<br>N | MRI | Brain volume | ROI | N | Y | Y | Y | N | N | Y |
| Kwak et al.<br>(2022) | 541<br>72.9<br>N | Y (*GDS +*<br>*NPI*)<br>Y (*NPI*) | Y<br>Y | MRI | Cortical thickness | ROI | N | N | N | N | N | N | Y |
| Özel et al.<br>(2022) | 4943<br>64.6<br>N | Y (*CES-D*)<br>Y (*M-CIDI*) | Y<br>N | MRI | Brain volume, FA and<br>MD | Whole<br>brain | Y | N | N | N | N | N | Y |
| Jones et al.<br>(2022) | 63<br>? | N<br>Y (*BAI*) | N<br>Y | MRI | Grey matter volume | ROI | N | N | N | N | N | N | N |
|  | N |  |  |  |  |  |  |  |  |  |  |  |  |
| Valsdottir et al. (2022) | 1707<br>74.24<br>N | Y (GDS)<br>N | Y<br>N | MRI | Relative grey matter volume and white matter volume | Whole brain | N | N | N | N | N | N | Y |
| Calarco et al. (2022) | 48<br>69<br>Y | Y (MADRS + PHQ-9)<br>N | Y<br>N | MRI | Locus coeruleus integrity | ROI | N | N | N | N | N | N | N |
| Zhang et al. (2022) | 7844<br>?<br>N | Y (PHQ-2)<br>N | Y<br>N | Both | Brain volume (total, white and grey), FA, intracellular volume fraction, ISOVF, orientation dispersion | Whole brain | N | N | N | N | N | N | Y |
| Petkus et al. (2022) | 764<br>81.6<br>N | Y (GDS)<br>N | Y<br>N | MRI | Grey matter volume | ROI | N | N | N | N | N | N | Y |
| Yu et al. (2021) | 91<br>71.3<br>N | Y (GDS)<br>Y (GAI) | Y<br>Y | DTI | Structural connectivity | Whole brain | N | N | N | N | N | N | Y |
| Manning et al. (2021) | 160<br>58.54<br>Y | Y (MADRS)<br>Y (STAI-S) | Y<br>Y | MRI | Brain volume and white matter lesions | ROI | N | Y | N | N | N | N | N |
| He et al. (2021) | 71<br>69.4<br>N | Y (DSM-IV + HAM-D)<br>N | Y<br>N | DTI | FA, MD, AD and RD | Both | N | Y | N | N | N | N | Y |
| Zhang et al. (2020) | 1188<br>78.11<br>N | Y (GDS-15)<br>Y (NPI-Q) | Y<br>N | MRI | Brain volume | ROI | Y | Y | N | N | N | N | Y |
| Christman et al. (2020) | 324<br>49.21<br>Y | Y (MADRS + DSM-IV-TR ) | Y<br>N | MRI | Brain volume (to determine brain age gap), white matter | ROI | N | N | N | Y | Y | N | N |
|  |  | N |  |  | hyperintensity volume |  |  |  |  |  |  |  |  |
| Rashidi-Ranjbar et al. (2020) | 333<br>71.4<br>Y | Y (DSM-IV)<br>N | N<br>N | Both | FA, MD, subcortical volume and cortical thickness | ROI | N | Y | Y | Y | N | N | N |
| Wu et al. (2020) | 70<br>65.43<br>Y | Y (HAMD)<br>N | Y<br>N | DTI | White matter networks | ROI | N | Y | N | N | N | N | Y |
| Li et al. (2020) | 389<br>65.5<br>N | Y (BDI)<br>N | Y<br>N | DTI | FA, MD, AD and RD | Whole brain | N | N | N | N | N | N | Y |
| Neth et al. (2020) | 537<br>58.7<br>N | Y (BDI)<br>N | Y<br>N | Both | FA, AD signature cortical thickness and intracranial volume | ROI | N | N | N | N | N | N | Y |
| Szymkowicz et al. (2019) | 80<br>70.7<br>N | Y (BDI-II)<br>N | Y<br>N | MRI | Grey matter volume | ROI | N | Y | N | N | N | N | Y |
| Kim et al. (2019) | 65<br>72.86<br>N | Y (GDS)<br>N | Y<br>N | MRI | Grey matter volume | ROI | N | N | N | N | N | N | Y |
| Zotcheva et al. (2019) | 751<br>58.9<br>N | Y (HADS-D)<br>Y (HADS-A) | Y<br>Y | MRI | Brain volume and brain parenchymal fraction | ROI | N | N | N | N | N | N | Y |
| Zeki Al Hazzouri et al. (2018) | 1111<br>70.82<br>N | Y (CES-D)<br>N | Y<br>N | MRI | White matter hyperintensity volume, TIV, TCV, hippocampal volume | ROI | N | Y | N | N | N | N | Y |
| Sin et al. (2018) | 52<br>67.92<br>Y | Y (HAMD)<br>N | Y<br>N | MRI | Grey matter volume | Whole brain | N | Y | Y | Y | N | N | N |
| Andreescu | 59 | Y (HAMD) | Y | Both | Cortical thickness, | ROI | N | Y | N | Y | Y | N | N |
| et al. (2017) | 68.73<br>Y | Y (*HAMA + GADSS*) | N |  | grey matter volume, MD, WMH burden and FA |  |  |  |  |  |  |  |  |
| Szymkowicz et al. (2017) | 73<br>71.79<br>N | Y (*BDI-II*)<br>N | Y<br>N | MRI | Cortical thickness and surface area | ROI | N | Y | N | N | N | N | Y |
| Emsell et al. (2017) | 107<br>73.16<br>Y | Y (*DSM-IV + GDS + MADRS*)<br>N | Y<br>N | Both | Apparent fibre density, cortical thickness, surface area, FA, RD, WMH volume, whole brain volume | ROI | N | N | N | Y | N | N | Y |
| McLaren et al. (2017) | 43<br>69.45<br>N | Y (*CES-D*)<br>N | Y<br>N | MRI | Grey matter volume | Whole brain | N | N | N | N | N | N | Y |
| Hermesdorf et al. (2017) | 636<br>50.53<br>Y | Y (*CES-D*)<br>N | Y<br>N | DTI | FA | Both | Y | N | N | N | N | N | N |
| Rousotte et al. (2017) | 738<br>75.5<br>N | Y (*GDS*)<br>N | Y<br>N | MRI | Grey matter volume | ROI | N | N | N | N | N | N | Y |
| Hybels et al. (2016) | 381<br>70.1<br>Y | Y (*DDES (including CES-D) + DIS*)<br>N | Y<br>N | MRI | White matter hyperintensity volume | ROI | N | N | N | N | N | N | Y |
| Szymkowicz et al. (2016) | 43<br>68.8<br>N | Y (*CES-D*)<br>N | Y<br>N | MRI | Cortical thickness | Both | N | Y | N | N | N | N | Y |
| Jayaweera et al. (2016) | 111<br>64.13 | Y (*DSM-IV + HAMD*) | Y<br>N | MRI | Grey matter volume | ROI | N | Y | Y | Y | N | N | Y |
|  | Y | N |  |  |  |  |  |  |  |  |  |  |  |
| Potvin et al. (2015) | 393<br>73<br>N | Y (CES-D)<br>Y (STAI) | Y<br>Y | MRI | Cortical thickness | ROI | N | N | N | N | N | N | Y |
| Charlton et al. (2015) | 76<br>67.47<br>Y | Y (HAMD + DSM-IV)<br>N | Y<br>N | DTI | White and grey matter networks | ROI | N | Y | N | N | N | N | N |
| Elcombe et al. (2015) | 118<br>67.3<br>N | Y (DSM-IV + GDS-15)<br>N | Y<br>N | MRI | Brain volume | ROI | N | Y | Y | Y | N | N | Y |
| Jayaweera et al. (2015) | 99<br>?<br>Y | Y (HAMD)<br>N | Y<br>N | MRI | Grey matter volume | ROI | Y | Y | Y | Y | Y | Y | N |
| Kirton et al. (2014) | 116<br>68.78<br>N | Y (CES-D)<br>N | Y<br>N | MRI | White matter lesion volume | ROI | N | N | N | N | N | N | Y |
| Tudorascu et al. (2014) | 314<br>73.18<br>N | Y (CES-D)<br>N | Y<br>N | Both | Grey matter volume, white matter hyperintensities and FA | Whole brain | N | N | N | N | N | N | Y |
| Hybels et al. (2014) | 377<br>69.8<br>Y | Y (DDES (including CES-D) + DSM-IV)<br>N | Y<br>N | MRI | White matter hyperintensity volume | ROI | N | Y | N | N | N | N | Y |
| Shimada et al. (2014) | 4352<br>72<br>N | Y (GDS-15)<br>N | Y<br>N | MRI | Grey matter volume | ROI | N | N | N | N | N | N | Y |
| de Diego-Adelino et al. (2014) | 69<br>45.73<br>Y | Y (HAMD)<br>N | Y<br>N | DTI | FA | Both | N | Y | Y | Y | Y | Y | Y |
| Sachs-Ericsson et<br>al. (2014) | 246<br>69.51<br>Y | Y (*DDES +*<br>*MADRS*)<br>N | Y<br>N | MRI | White matter lesions | Whole<br>brain | N | N | Y | Y | N | N | Y |
| Ezzati et al.<br>(2013) | 36<br>82.2<br>N | Y (*GDS*)<br>Y (*SF-36*) | Y<br>Y | MRI | Grey matter volume | ROI | Y | Y | N | N | N | N | Y |
| Zannas et<br>al. (2013) | 159<br>69.56<br>Y | Y (*MADRS*)<br>N | Y<br>N | MRI | Grey matter volume | ROI | N | N | N | N | N | N | N |
| Sachs-Ericsson et<br>al. (2013) | 135<br>70.55<br>Y | Y (*MADRS*)<br>N | Y<br>N | MRI | Grey matter volume | ROI | N | N | N | Y | N | N | Y |
| Lee et al.<br>(2012) | 161<br>74.4<br>N | Y (*DSM-*<br>*IIIR*)<br>N | Y<br>N | MRI | White matter lesion<br>volume | ROI | N | N | N | N | N | N | Y |
| Lim et al.<br>(2012) | 95<br>71.26<br>Y | Y (*HAMD*)<br>N | Y<br>N | MRI | Cortical thickness<br>and subcortical<br>volumes | Whole<br>brain | N | Y | N | Y | Y | N | Y |
| Steffens et<br>al. (2011) | 162<br>69.68<br>Y | Y (*HAMD +*<br>*MADRS +*<br>*CGIS +*<br>*DDES*)<br>N | Y<br>N | MRI | Grey matter volume | ROI | N | Y | N | Y | N | N | Y |
| Kohler et al.<br>(2010) | 64<br>73.51<br>Y | Y (*MADRS*<br>*+ DSM-IV*)<br>N | Y<br>N | MRI | Brain volume + white<br>matter<br>hyperintensity<br>severity | ROI | N | Y | Y | Y | Y | Y | N |
| Kieseppa et<br>al. (2010) | 36<br>44.84<br>Y | Y (*BDI +*<br>*DSM-IV*)<br>Y (*BAI +*<br>*DSM-IV*) | Y<br>N | DTI | FA | Whole<br>brain | N | N | Y | N | Y | N | N |
| Lamar et al. (2010) | 79<br>68.6<br>N | Y (GDS)<br>N | Y<br>N | Both | FA, MD and white matter hyperintensities | ROI | N | N | N | N | N | N | Y |
| Ries et al. (2009) | 47<br>67.73<br>N | Y (CES-D)<br>N | Y<br>N | MRI | Grey matter volume | Whole brain | N | N | N | N | N | N | Y |
| Dotson et al. (2009) | 110<br>69.38<br>N | Y (CES-D)<br>N | Y<br>N | MRI | Brain volume | ROI | N | Y | N | N | N | N | Y |
| Lavretsky et al. (2008) | 270<br>74.4<br>N | Y (MUDS)<br>N | Y<br>N | MRI | Grey and white matter volumes, lacunar volumes in subcortical white and grey matter structures, volume of white matter hyperintensities and total hippocampal volume. | ROI | N | N | N | N | N | N | Y |
| Potter et al. (2007) | 130<br>72.49<br>Y | Y (MADRS + DSM-IV)<br>Y (DSM-IV) | Y<br>Y | MRI | White matter lesion volume | ROI | Y | Y | N | N | N | N | N |
| Sachdev et al. (2005) | 412<br>62.54<br>N | Y (PHQ-9)<br>Y (scale not given) | Y<br>N | MRI | Brain atrophy, mid-ventricular VBR, periventricular white matter hyperintensities/total white matter and deep white matter hyperintensities/total white matter | Both | N | Y | N | N | N | N | Y |
| Neu et al.<br>(2005) | 57<br>53.04<br>Y | Y (*DSM-IV*<br>*SCIDI*)<br>N | Y<br>N | MRI | White matter<br>hyperintensities | Whole<br>brain | N | Y | Y | Y | N | N | N |
| Janssen et<br>al. (2004) | 69<br>63.05<br>Y | Y (*DSM-IV*<br>*MINI* +<br>*MADRS*)<br>N | Y<br>N | MRI | Grey and white<br>matter volumes | ROI | N | Y | N | Y | Y | N | Y |
| Almeida et<br>al. (2003) | 88<br>73.67<br>Y | Y (*MADRS*<br>+ *DSM-IV*)<br>N | Y<br>N | MRI | Brain volume | ROI | N | N | N | Y | N | N | Y |
| Nebes et al.<br>(2002) | 13<br>73.54<br>Y | Y (*HAMD* +<br>*DSM-IV*)<br>N | Y<br>N | MRI | White matter<br>hyperintensity<br>volume | ROI | N | N | Y | N | N | N | Y |
| Wohl et al.<br>(1994) | 16<br>63<br>N | Y (*DS-II-R*<br>*SCIDI* +<br>*HAMD*)<br>N | Y<br>N | MRI | White matter<br>hyperintensities | Whole<br>brain | N | N | N | N | N | N | Y |

We excluded 32 papers at the full screen stage with the reasons for exclusion being coded. There were four reasons for exclusion: not measuring the variables of interest; participants not within the agreed age range); wrong study design; and conference abstracts (where there was no full paper).

We applied an additional level of exclusion where any studies that lacked a clear description of the direction of the effect or did not include information about the direct effect between depression or anxiety and brain structure were removed. This resulted in 3 additional papers being removed (Cotter et al., 2020; Kim & Kim., 2022; Sun et al., 2023).

From each of the included studies, data was extracted including but not limited to: the study characteristics, the publication year, the databases searched, the study population and setting, the outcome measures and covariates controlled for. An Excel spreadsheet was created for data extraction purposes.

### Participants

In the final 62 papers selected, the number of participants varied widely (Ns were minimum 13, maximum 7844, mean 539.56, SD 1276.75).

The age of participants also varied widely. Mean ages ranged from 44.84-82.2 years.

29 papers (46.77%) used clinical participants, meaning participants with a diagnosis or who met diagnostic criteria for depression or anxiety. The remaining papers assessed depression or anxiety symptoms in “healthy” middle-aged or older adults. This means that the population was not selected for their depression status, but some participants did meet criteria for depression or anxiety when assessed.

### Depression and Anxiety Measures

There were considerably more papers examining depression than anxiety. 50 papers (78.46%) measured depression, 11 measured both (16.92%) and only one paper measured anxiety on its own. However, of the 11 papers that measured both, three of these (27.27%%) did not include anxiety in their brain analysis.

Despite the fact that depression and anxiety are highly comorbid, only five papers (8.06%%) controlled for the other disorder when examining depression or anxiety.

Numerous depression and anxiety measurement tools were used, which likely reflects the fact that differing psychological measures are appropriate for different situations or populations. For example, different measures may be used for measuring current or long-term symptoms and there are specific scales for certain populations, such as the Geriatric Depression Scale (GDS, Scogin et al., 2000). Overall, there are 23 unique measures employed by the papers included in this review. There were six different measures used to measure anxiety, 15 to measure depression and two that measured both anxiety and depression. The most common depression measures were The Structured Clinical Interview from the Diagnostic and Statistical Manual of Mental Disorders (DSM-IV-SCI, American Psychiatric Association, 1994), used in 26.23%% of papers that measured depression, the Hamilton Depression Scale, (HAM-D, Hamilton, 1960), used in 21.31% of papers that measured depression the Geriatric Depression Scale (GDS, Scogin et al. 2000), and the Centre for Epidemiological Studies Depression Scale (CES-D, Radloff, 1977) both also used in 20.96% of the papers that measured depression. The most common anxiety measures were the State-Trait Anxiety Inventory (STAI, Spielberger, 1989), the Neuropsychiatric Inventory (NPI, Cummings, 1997), the Beck Anxiety Inventory (BAI, Beck et al., 1988) and the Structured Clinical Interview from the Diagnostic and Statistical Manual of Mental Disorders (DSM-IV-SCI, American Psychiatric Association, 1994) all used in 16.67%% of papers that measured anxiety, however, this is only two papers each).

### Neuroimaging Variables

Of the final 62 papers, 46 (74.19%) used T1 MRI, 8 (13.33%) used DTI and 8 (13.33%) used both. There were several brain measures including total brain volume, grey matter volume, cortical thickness, fractional anisotropy (FA), mean diffusivity (MD) and white matter lesion volume amongst others (see Table 1).

### Direction of Effects

Of the 46 studies finding significant correlates of depression, 37 (80.43%) of these had a negative relationship, such that increased depressive symptom scores were associated with diminished brain structure, such as reduced grey matter volume, reduced cortical thickness, or increased white matter hyperintensity/lesion volume. Three papers found positive relationships and six papers found both positive and negative relationships between depression and brain structure which was region specific. For the two studies finding significant correlates of anxiety and brain structure, both of these found negative relationships.

There were three papers that found significant correlates of both depression and anxiety and brain structure. Of these, one found a negative relationship and two papers found both positive and negative relationships.

Despite the overwhelming majority of studies reporting negative effects, some papers did find positive effects, indeed sometimes in the same brain regions as negative effects were found by others (See Table 2 and 3).

**Table 2:**
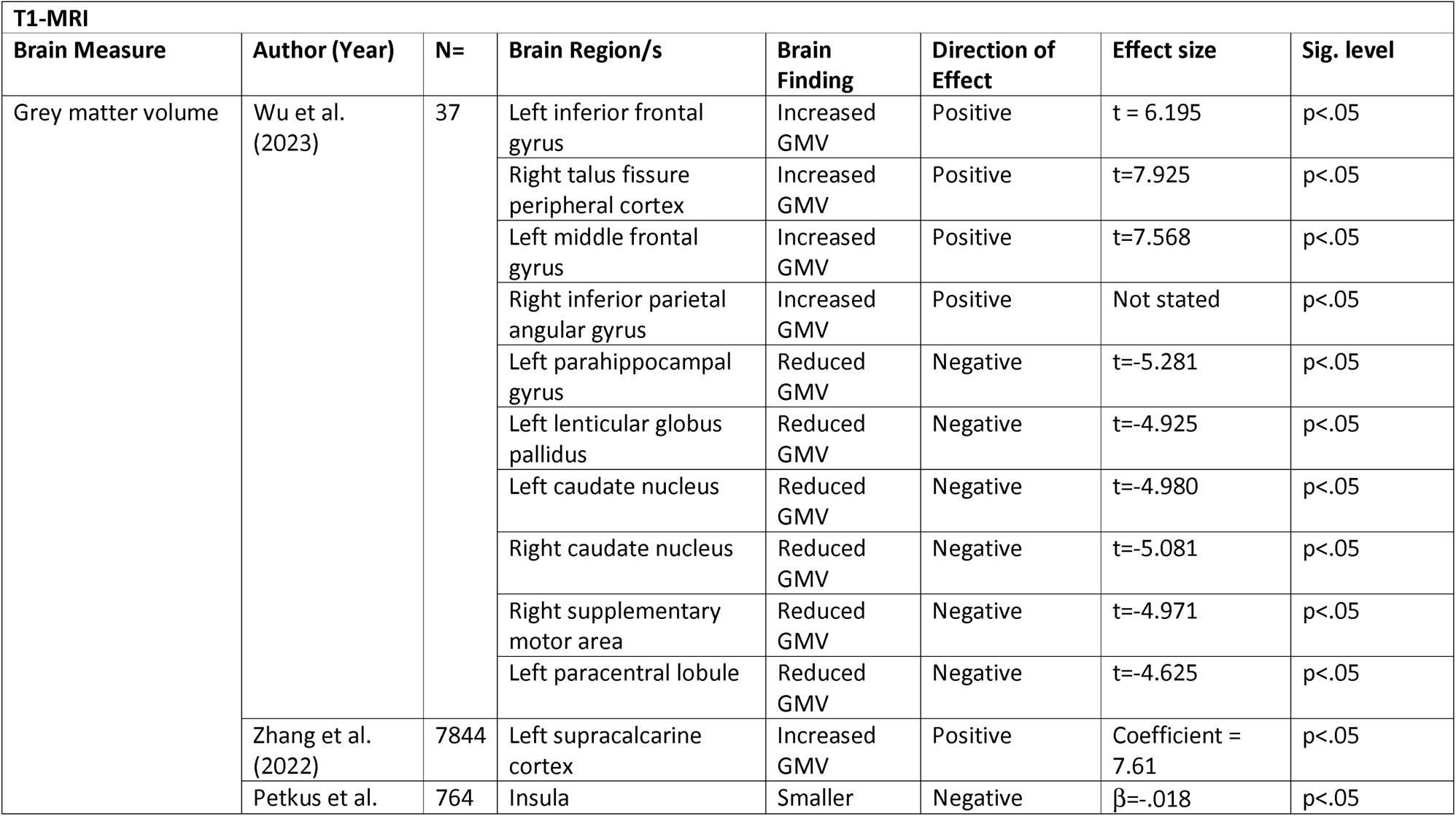
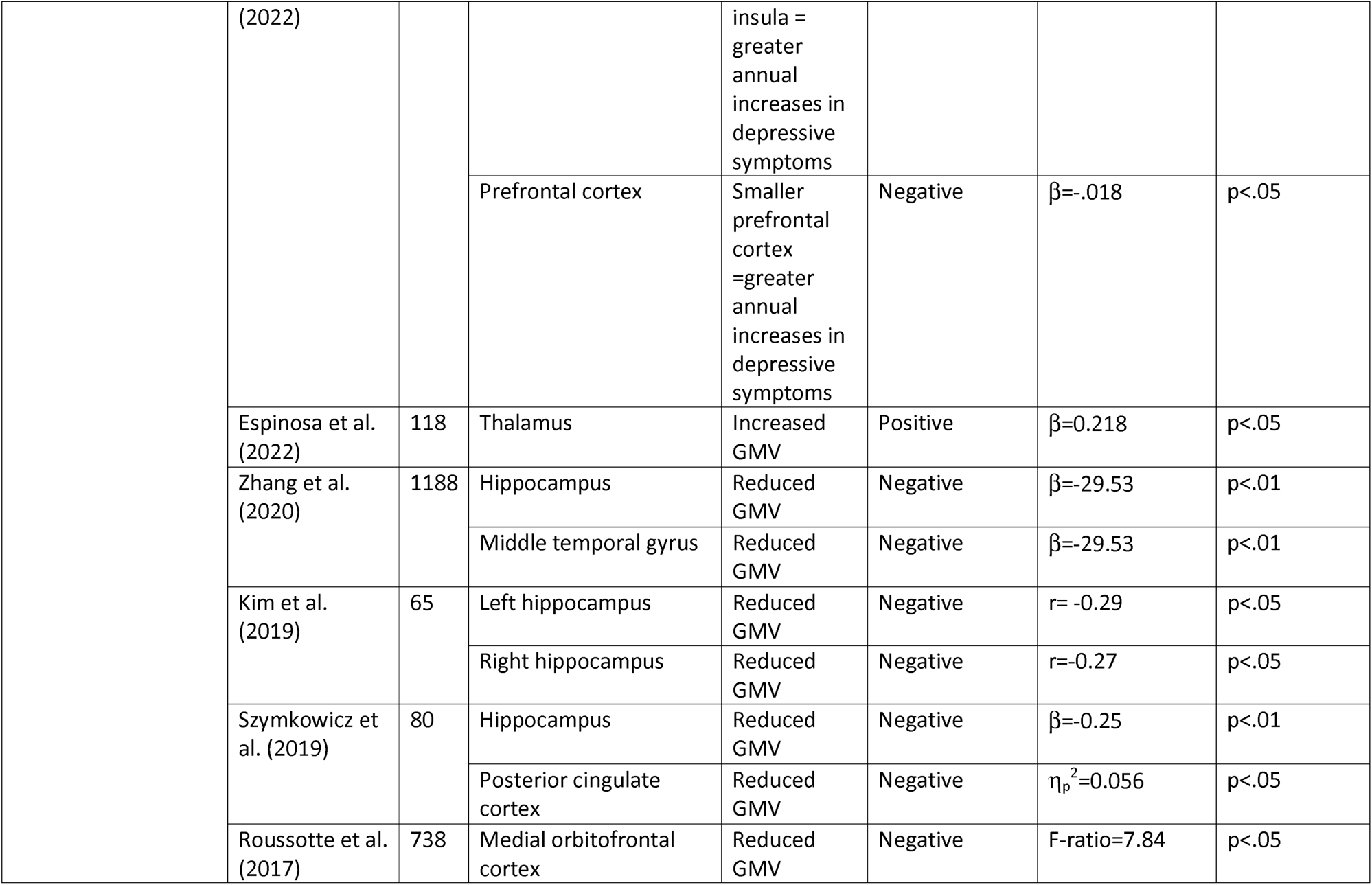
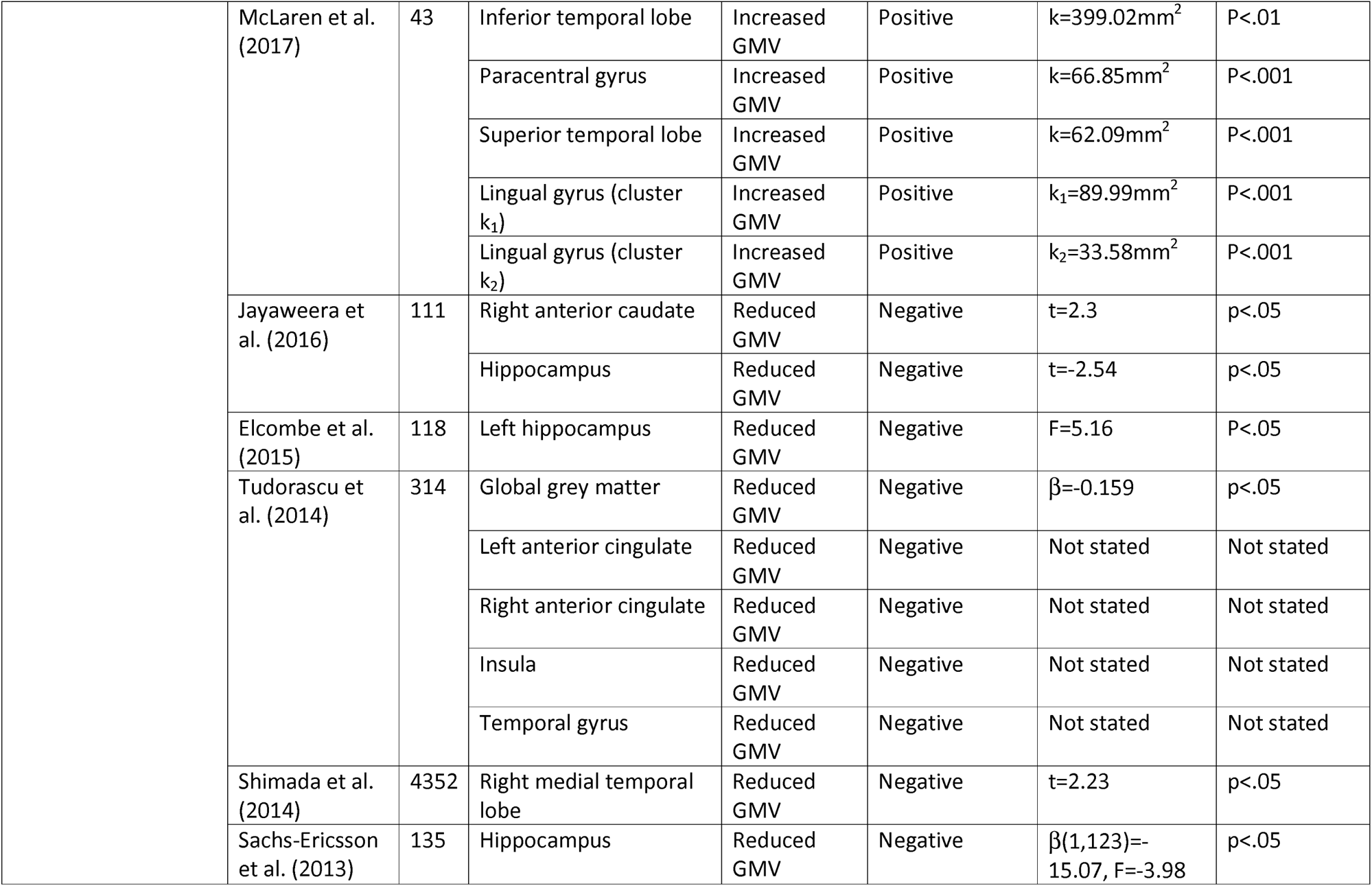
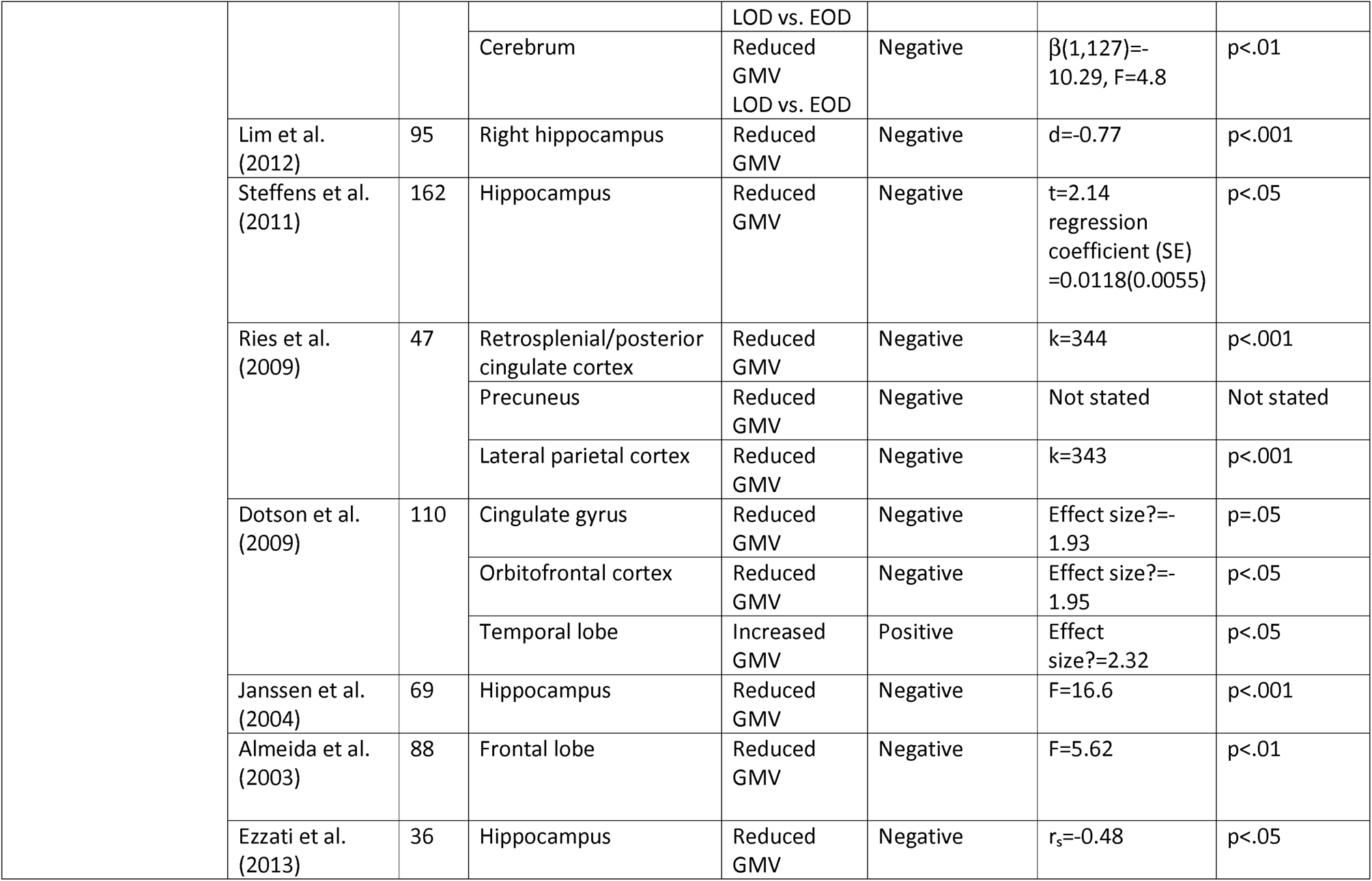
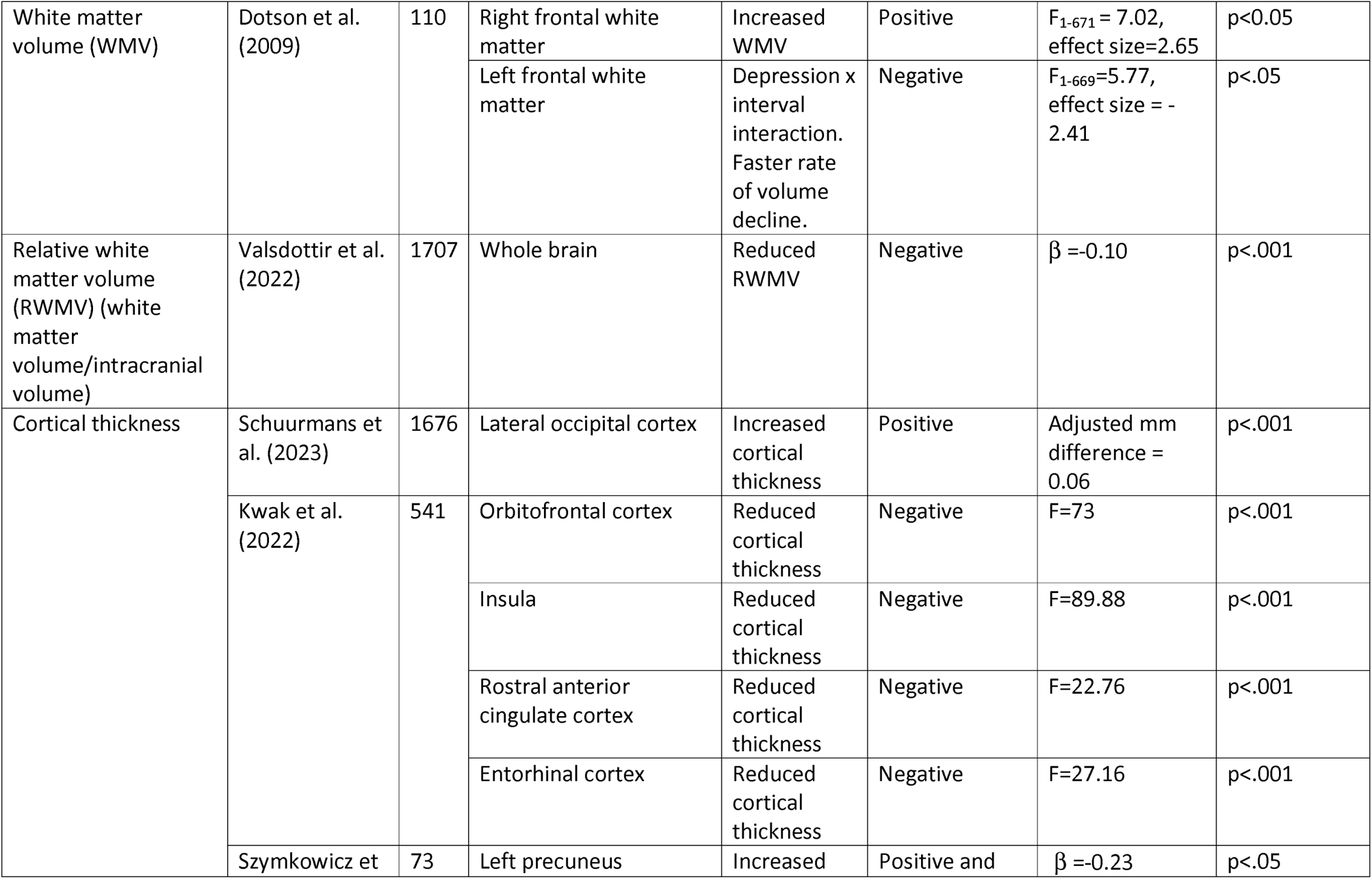
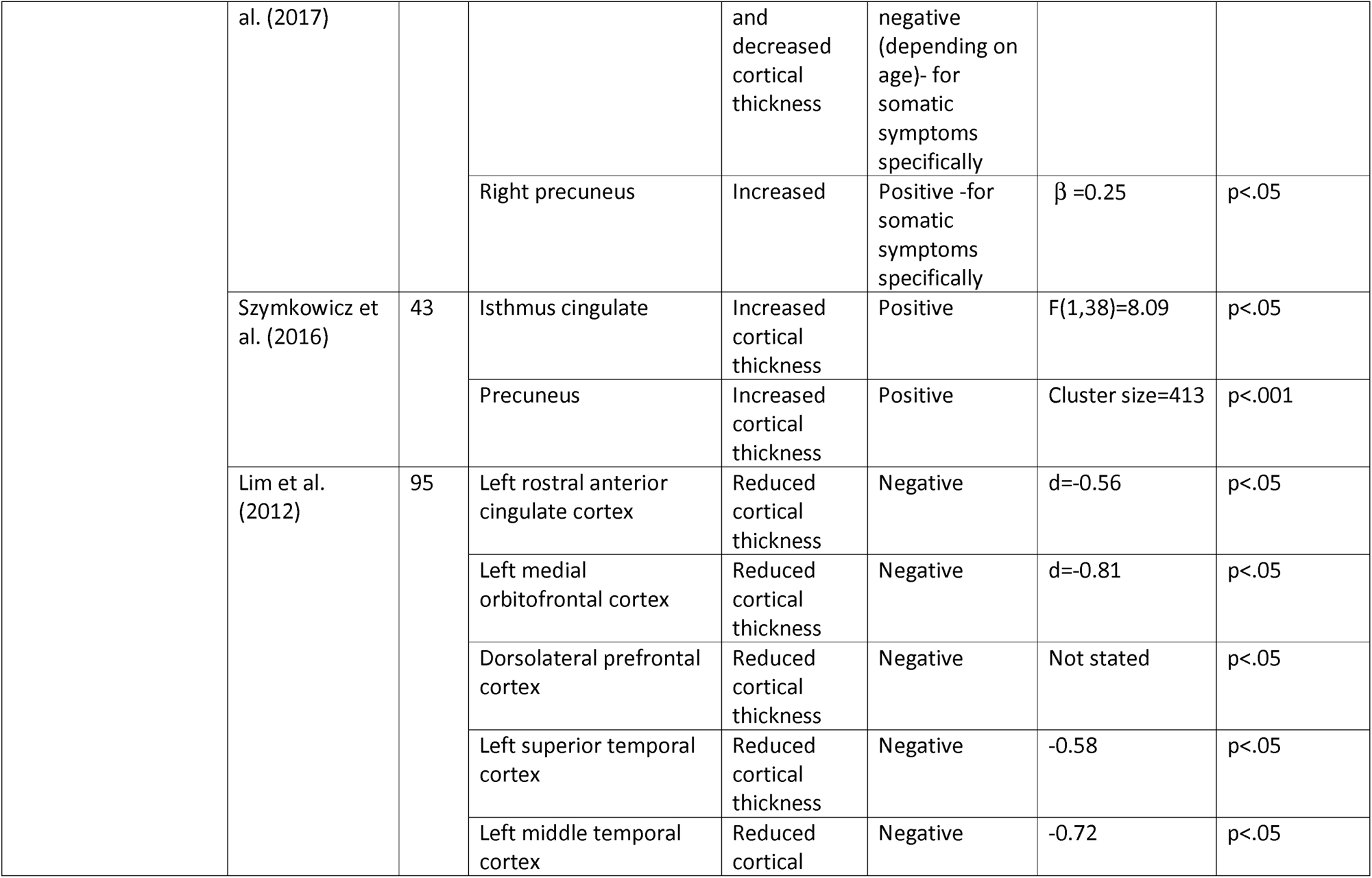
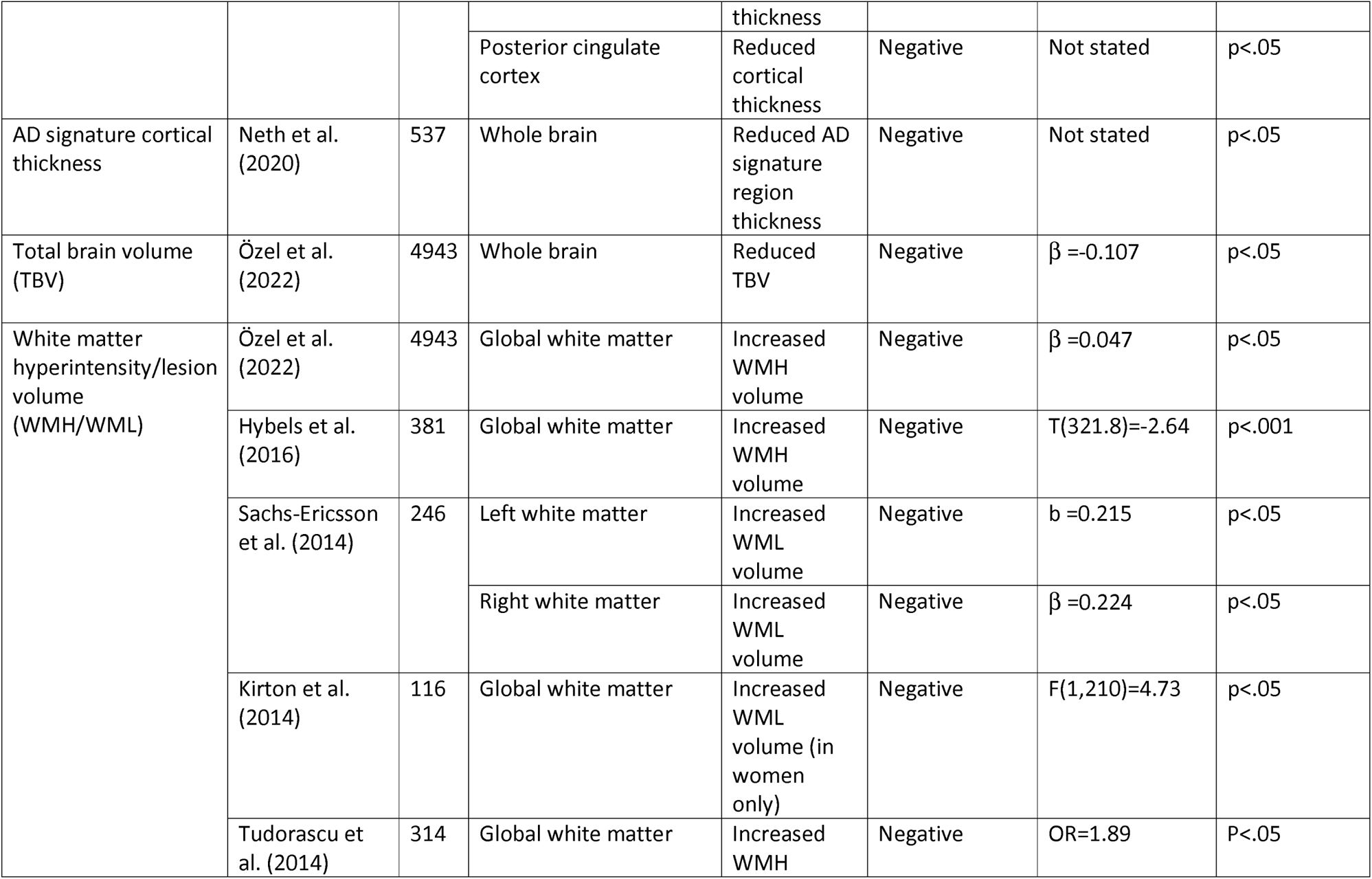
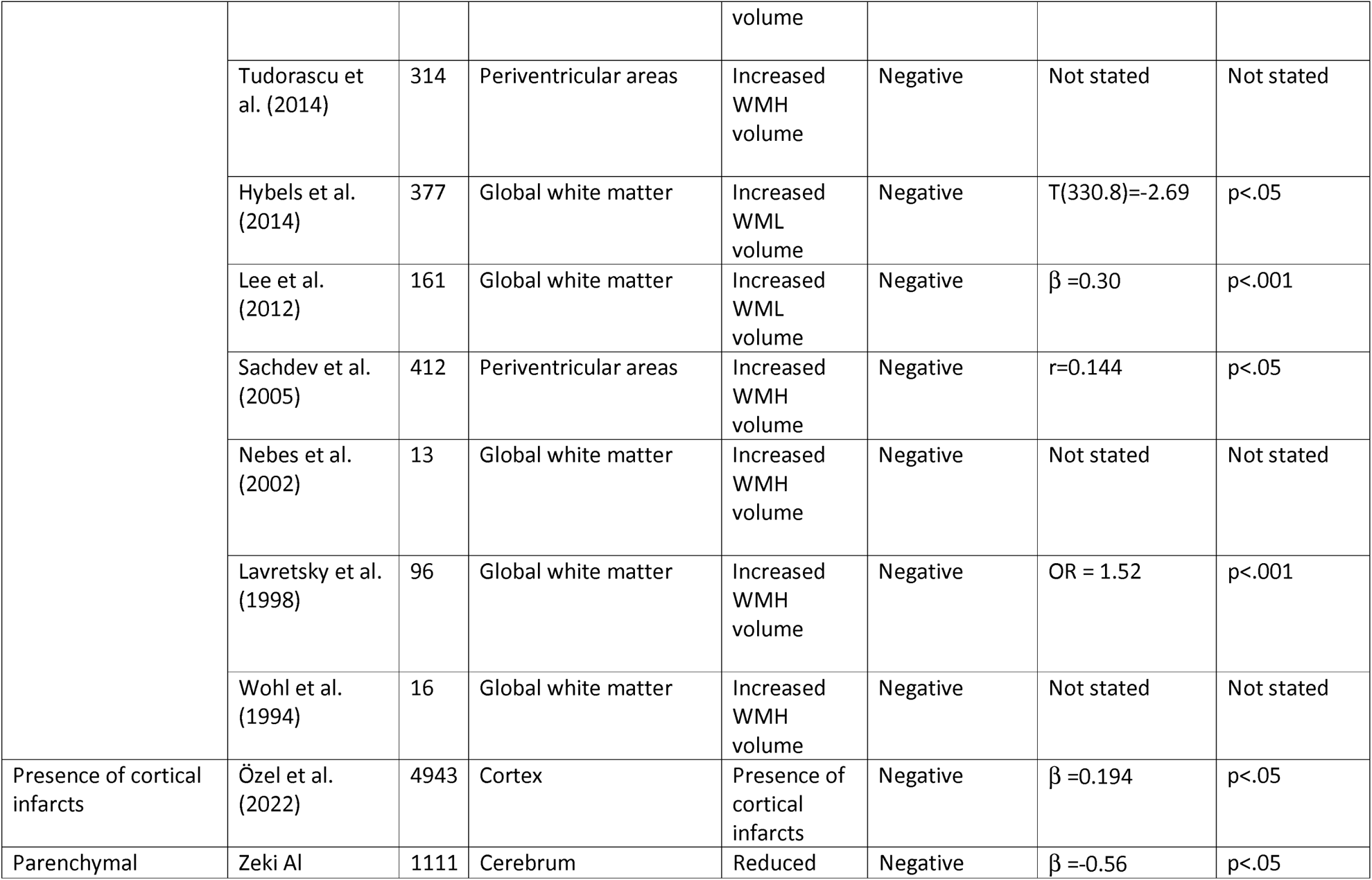
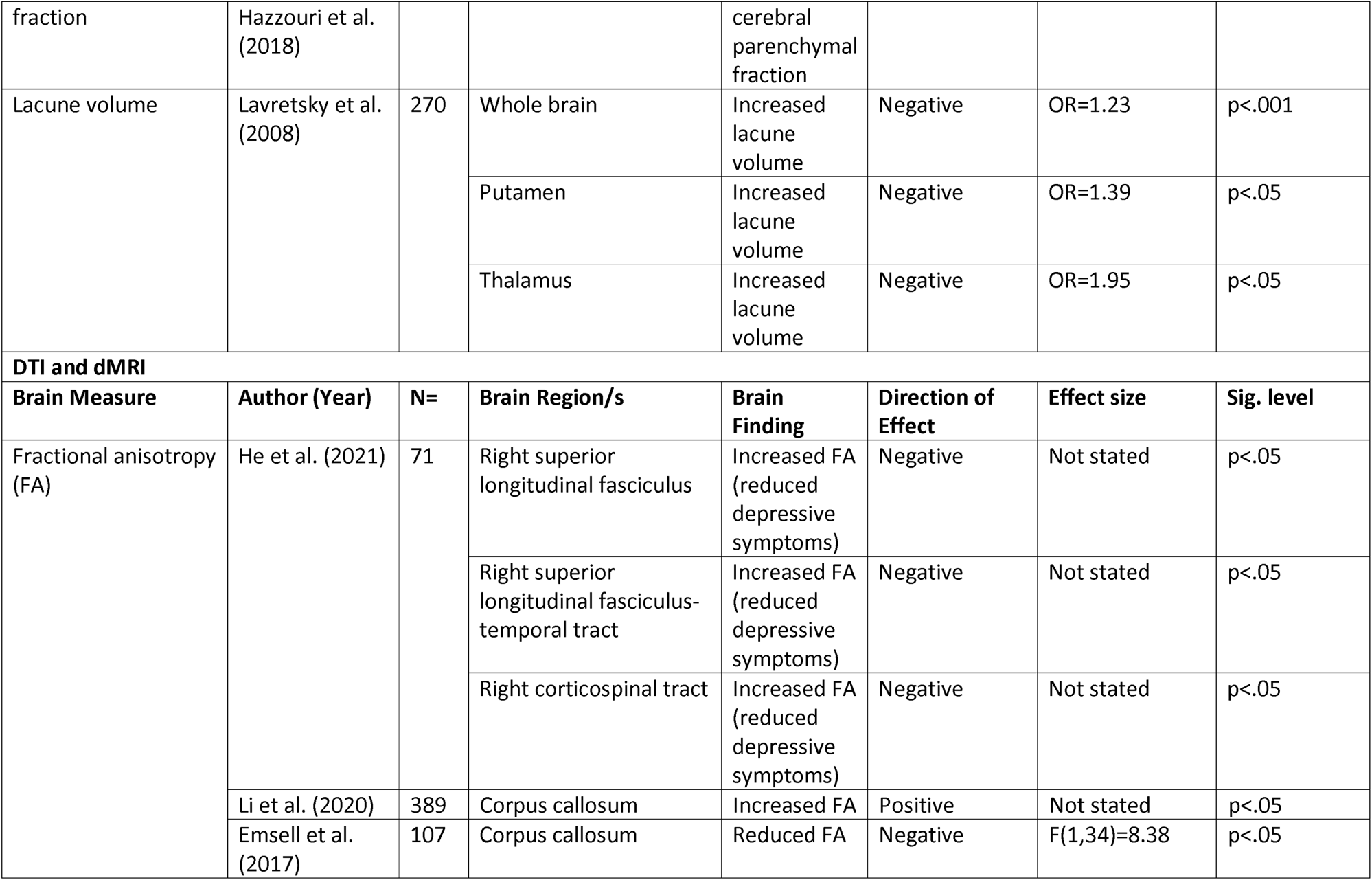
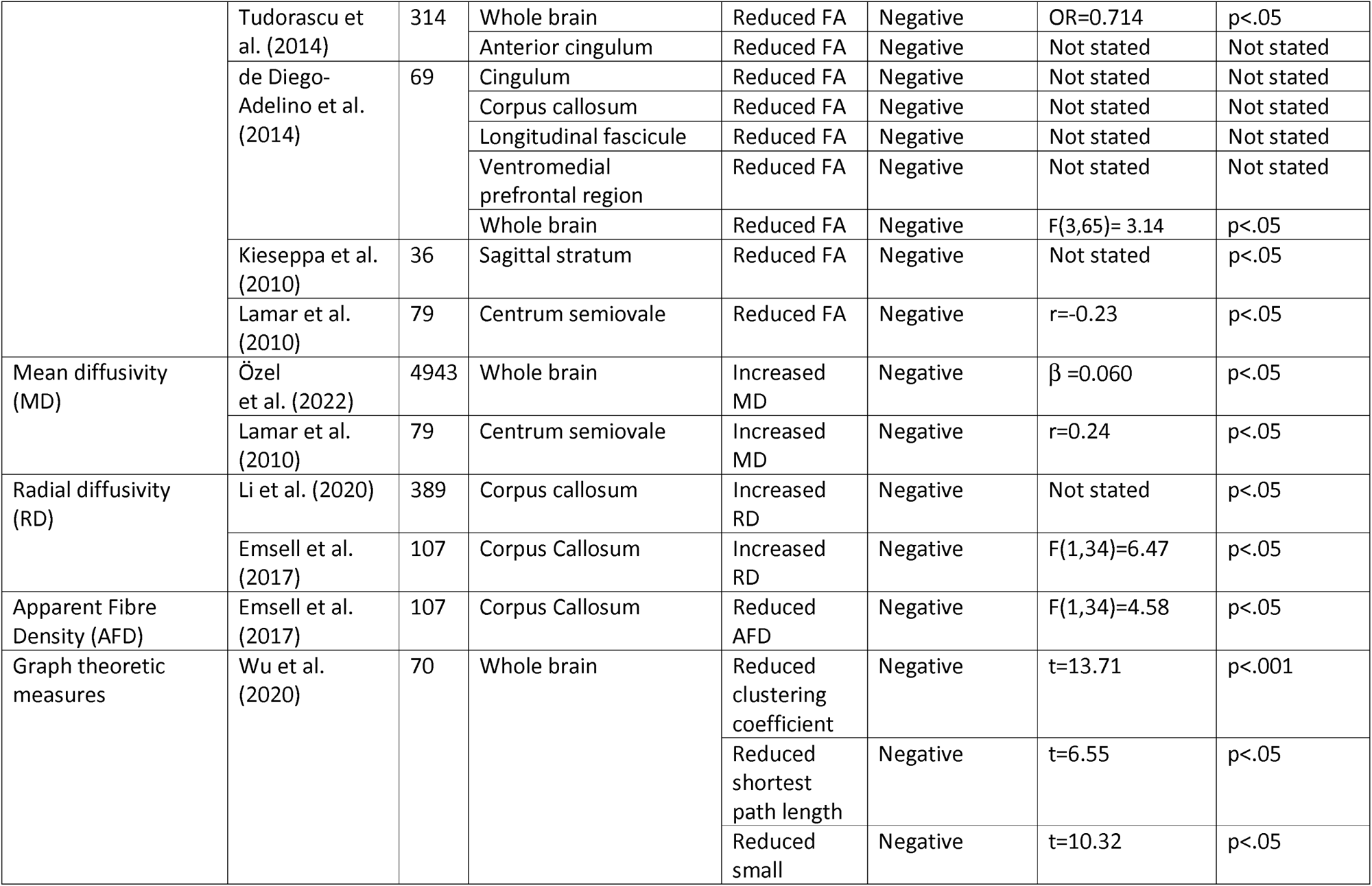
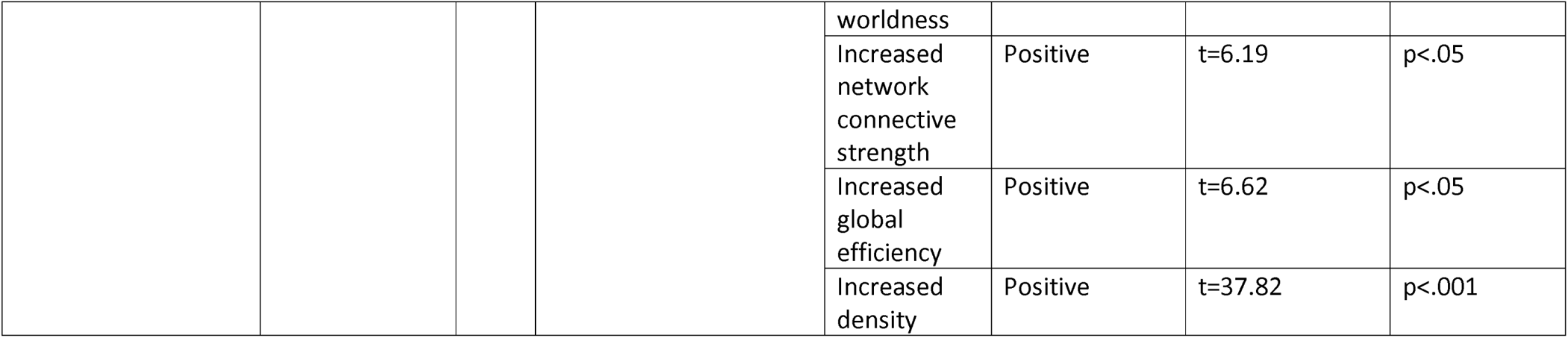
Brain regions implicated in depression.

**Table 3:**
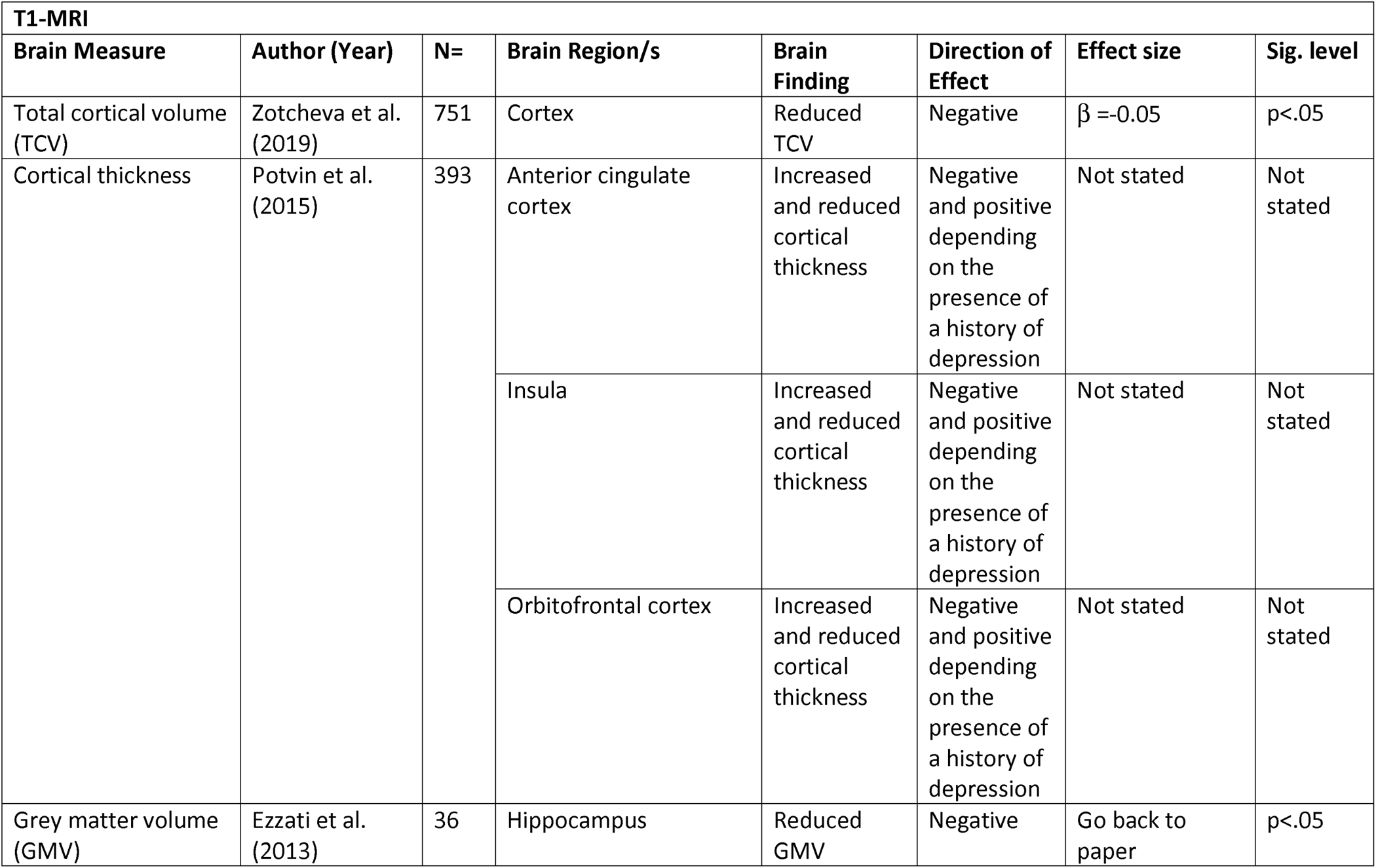
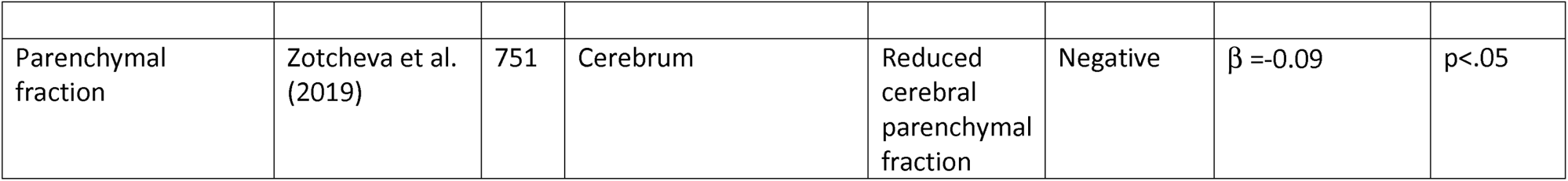
Brain regions implicated in anxiety.

### Whole Brain vs. ROI Analyses

The majority of studies (41 (66.13%)) used ROI analysis, whereas 16 (25.81%) used whole brain and 5 (8.06%) conducted both.

### Consideration or Controlling of Covariates

There are several variables that are known to have an impact on the relationship between depression and anxiety and brain structure. Despite this, several papers did not consider all or some of these variables.

#### Medication

Of the 62 papers in the final review, only 28 (45.16%) considered the effects of psychiatric medication on this relationship. Of the 29 papers that used clinical participants, 19 (65.52%) considered the effect of psychiatric medication on this relationship.

#### Number of episodes

Only 13 (20.96%) papers considered the number of episodes of depression or anxiety that the participants in the study had experienced. This is not particularly surprising given that many of the studies examined depression and anxiety in “healthy” adults that did not necessarily have a diagnosis of depression. Of the 29 papers that used clinical participants, 12 (41.38%) considered the impact of the number of episodes experienced.

#### Age at onset

Only 19 (30.64%) papers reported the age of onset of depression or anxiety. As above, if an individual did not have a diagnosis of depression, then this may not be possible to determine. However, we know it can impact the relationship studied. Of the 29 papers that used clinical participants, 18 (62.07%) considered the effect of age of onset on this relationship.

#### Duration of symptoms

Only 9 (14.52%) of papers considered the duration of symptoms of depression or anxiety. Again, in a situation with a lack of diagnosis, this information may be difficult to collect. However, of the 29 papers that used clinical participants, and thus could collect the information, only 8 (27.59%) considered duration of symptoms.

#### All of the above

Only three (4.84%) of the papers in the final review considered all of these variables. Of the 29 papers that used clinical participants, only three (10.34%) considered all of these variables.

### Consideration of Social Factors

Of the 55 papers (88.71%) that measured at least one social factor, most commonly education, only eight (14.54%) discussed the effect that these social factors could have on depression or anxiety and brain structure or the relationships between them. If social factors were measured, they were usually completely ignored in analyses or used simply as covariates.

### Sex Disaggregation

Despite many papers using sex as a covariate, none of the papers in the final review ran their analyses separately for males and females. This means that it is not possible to identify whether the relationships between depression or anxiety and brain structure differ between males and females.

### Structural Correlates

Since there was not a sufficient number of papers that used whole brain analysis, it is not possible to do a quantitative analysis to identify consistently reported brain regions associated with depression and/or anxiety. Nevertheless, some commonalities have been found across the literature, which are summarised for grey and white matter in turn below.

### Whole Brain Findings

Most of the studies (76.19%) that used whole brain analyses reported significant findings. The brain regions reported using whole brain analyses were somewhat varied without many papers reporting the same regions. However, the corpus callosum was implicated in two papers (Li et al., 2020; de Diego-Adelino et al., 2014). Both papers reported reduced FA associated with depressive symptoms with Li et al. (2020) in addition reporting increased RD, both of which indicate reduced structural integrity. The anterior cingulate cortex was also implicated in two papers, with Tudorascu et al. (2014) and Lim et al. (2012) both reporting reduced GMV in this region. Interestingly, Lim et al. (2012) also reported reduced GMV in the hippocampus and is the only paper using whole brain analyses to report a finding in the hippocampus, a region that is often the focus of depression and anxiety region and often the target in ROI analyses. Contradictory findings were reported in the precuneus with Szymkowicz et al. (2016) reporting increased depressive symptom severity being associated with greater cortical thickness and Ries et al. (2009) reporting reduced GMV in this region in depressed subjects relative to healthy controls. Three papers using whole brain analysis reported increased white matter hyperintensity/lesion volume in global white matter (Özel et al., 2022; Sachs-Ericsson et al., 2014; Wohl et al., 1994) and two reported increased white matter hyperintensity volume in periventricular areas (Tudorascu et al., 2014; Sachdev et al., 2005).

### Region of Interest (ROI) Findings

71.74% of papers using ROI analysis reported significant findings. Figure 2 shows the number of papers that had null, positive, and negative associations of depression or anxiety with specific regions of interest. Interestingly, the hippocampus, which is one of the main brain regions implicated across the papers in the review, showed almost as many null findings as significant ones. In addition, the amygdala, which is often cited as being related to emotions, had no significant findings despite being a region of interest in four studies. Temporal regions, which were examined as regions of interest in six studies, only showed significant findings in two studies, one with positive findings and one with negative.

**Figure 2:**
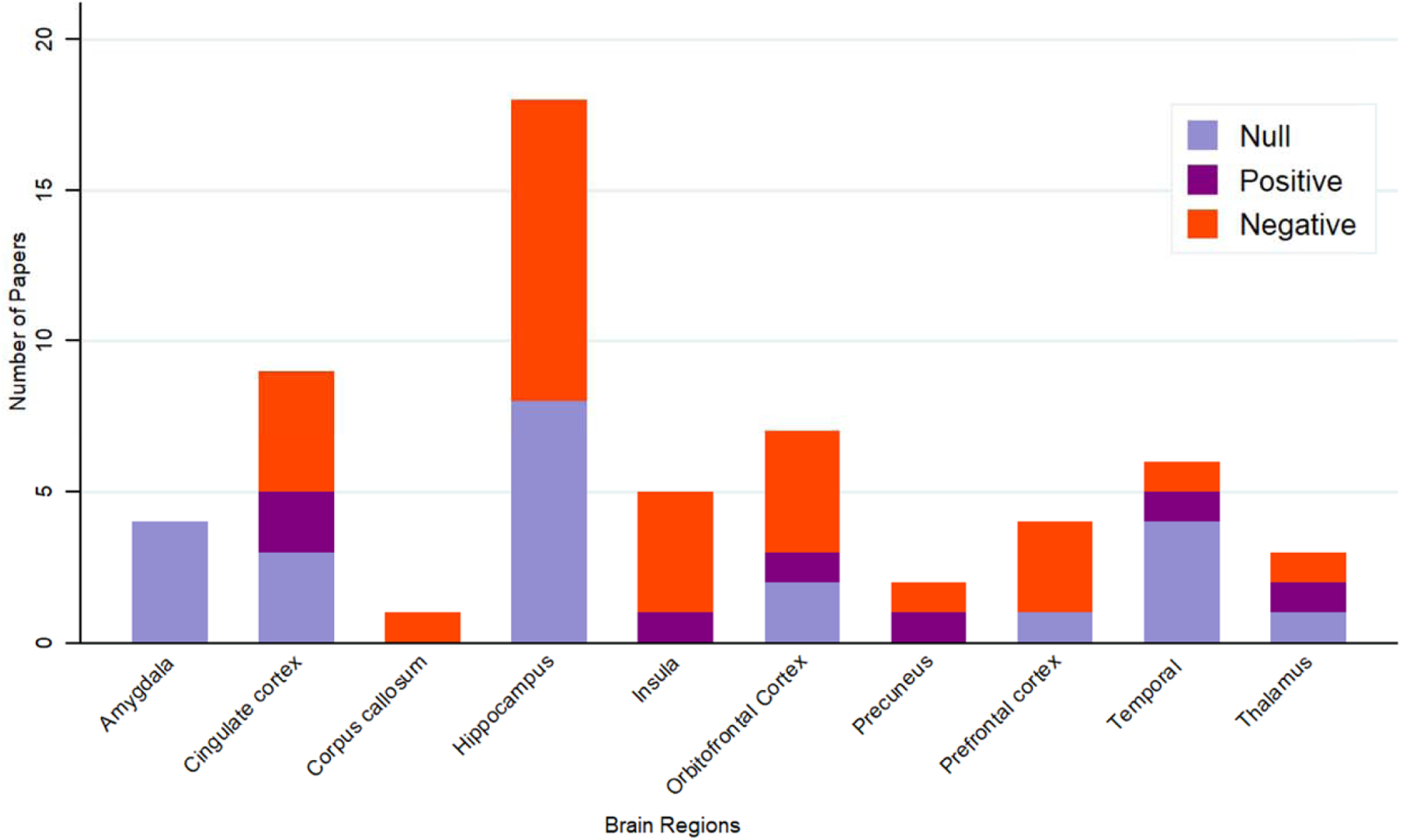
Association between depression/anxiety and brain regions in studies using ROI analysis

## DISCUSSION

With the current review, we aimed to establish whether there are any structural brain correlates that are consistently implicated in depression and anxiety in middle-aged and older adults. The areas that were reported most often for depression include the hippocampus, orbitofrontal cortex, prefrontal cortex, insula, temporal lobe/gyrus/cortex, the corpus callosum, and the cingulate cortex, both posterior and anterior. Given the small number of papers examining anxiety, it is not possible to say which areas are implicated with any certainty, however the anterior cingulate cortex (ACC) and orbitofrontal cortex were both implicated in two studies.

The hippocampus was implicated most often, which is unsurprising as it is thought to play a key role in the processing of emotions (Immordino-Yang & Singh, 2013; Femenía et al., 2012) and emotional memory (Perry et al., 2011). The relationship between severity of depression symptoms and structure of the hippocampus was consistently negative. Studies reported reduced GMV and cortical thinning in this area. It should be noted that nine out of the eleven studies that reported changes in the hippocampus used ROI analyses, and thus were specifically looking for findings in this area. Our review found mixed findings of the association between depression and brain structure in this region, while ten studies using ROI analyses found a significant negative relationship, eight papers using ROI analyses of the hippocampus failed to find a significant relationship. One paper also found reduced GMV in the hippocampus was associated with higher anxiety scores (Ezzati et al., 2013), this paper also used ROI analysis.

The cingulate cortex, both anterior and posterior and the cingulum were also implicated in multiple studies. Findings were reported in white and grey matter, and in both depression and anxiety. Most papers reported negative associations, however one paper (Potvin et al., 2015) reported an increase in cortical thickness in individuals who had trait anxiety without a past history of depression. They reported the opposite finding, reduced cortical thickness, in individuals with trait anxiety and a history of depression. This seems to further support the fact that anxiety and depression must both be considered in this area of research, as one’s presence can impact on the other’s relationship with brain structure. There were three studies that conducted ROI analyses of the cingulate cortex and did not find a significant relationship between its structure and depression or anxiety. Thus results in this region are inconsistent both in terms of whether a relationship is found and the direction of the relationship when it is found.

Changes to the structure of the orbitofrontal cortex were found to have a negative relationship with depression in four studies (Rousesotte et al., 2017; Dotson et al., 2009; Kwak et al., 2022; Lim et al., 2012). These studies reported both cortical thinning and reduced grey matter volume in this area in the presence of depressive symptoms. The orbitofrontal cortex was also found to be related to anxiety (Potvin et al. (2015). Potvin et al. (2015) reported increased cortical thickness in the orbitofrontal cortex of individuals with trait anxiety, without a history of depression. Again, they reported the opposite finding in this area in those who had a history of depression. All of the papers reporting orbitofrontal cortex findings in both depression and anxiety conducted ROI analyses.

Temporal regions including the temporal lobe, the temporal gyrus and the superior and middle temporal cortices were consistently associated with depressive symptoms. The majority of the relationships found in these areas were negative, specifically with reduced GMV or reduced cortical thickness (Zhang et al., 2020; Tudorascu et al., 2014; Shimada et al, 2014; Lim et al., 2012), however the inferior temporal lobe was positively associated with depression in McClaren et al’s (2017) paper. McLaren et al. (2017) demonstrated that larger GMVs in the left inferior temporal lobe were associated with higher depressed mood subscale scores. In addition, Dotson et al. (2009) found that higher depressive symptoms were associated with larger GMVs in left temporal regions.

Interestingly, while temporal regions were associated with depressive symptoms in many studies, in those that used ROI analyses of this areas there were four papers that did not find significant results, compared with just one paper finding a positive relationship and one finding a negative one.

The white matter of the prefrontal cortex was negatively associated with depression symptoms in two studies, and the ventromedial prefrontal region showed the same negative relationship in one study. Charlton et al. (2015) found that white matter vulnerability, a measure of how important a specific node is to the efficiency of the network, was significantly higher in the right prefrontal cortex in their late life depression group than the healthy older adult group. This means that, in those with late life depression, frontal regions, which have been implicated in mood regulation, are essential for the efficiency of the network (Charlton et al., 2015). Lim et al. (2012) investigated late onset depression (LOD) and found that compared to controls, individuals with LOD demonstrated reduced cortical thickness in the dorsolateral prefrontal cortex. de Diego-Adeliño et al. (2014) examined white-matter microstructure in patients at different stages of MDD and healthy controls. They found decreased FA in the ventromedial prefrontal region in treatment-resistant/chronic MDD, even when compared with the remitted-recurrent MDD group. Symptom severity and number of previous episodes were significant predictors of this relationship. Petkus et al. (2022) examined the grey matter of the prefrontal cortex and found that smaller prefrontal cortex was associated with greater annual increases in depressive symptoms. Taken altogether, there seems to be a decent amount of evidence of diminished brain structure in prefrontal areas relating to depression.

The insula was implicated in four papers. Three papers found significant negative relationships between brain structure and depression, and one found both a positive and negative relationship between brain structure and anxiety. Kwak et al. (2022) and Toradascu et al. (2014) found that depression was related to cortical thinning and reduced GMV respectively. As with grey matter in the prefrontal cortex, Petkus et al. (2022) found that a smaller insula was associated with greater annual increases in depressive symptoms. As with other regions discussed above, Potvin et al. (2015) found that individuals with trait anxiety without a history of depression had increased cortical thickness in the insula, whereas those with a history of depression had decreased cortical thickness. All studies finding a relationship in this region conducted ROI analyses and there were no studies that found null results in the insula.

The corpus callosum was associated with depression in five papers. It was associated with both increased (Li et al., 2020) and reduced (Emsell et al., 2017; de Diego-Adelino et al, 2014) FA, in addition to increased RD (Li et al., 2020; Emsell et al., 2017) and reduced apparent fibre density (AFD, Emsell et al., 2017). These associations were predominantly negative, meaning a reduction in structural integrity, as reduced FA and increased RD may indicate myelin loss or atrophy (Solowij et al., 2017; Masdeu & Pascual, 2016). As stated, however, a positive relationship was found by Li et al. (2020) in that depression scores were associated with increased FA, suggesting increased white matter integrity.

Thus, we have identified brain regions that are relatively consistently implicated in depression in middle-aged and older adults. The review has provided additional evidence for the fact that further research is needed into the relationship between anxiety disorders and brain structure in this population. Despite identifying certain candidate structural brain regions for depression in these age groups, the direction of effects is still not always consistent.

As with many of the regions most consistently implicated in depression and anxiety reported above, most papers in the review reported negative relationships between depression and anxiety symptoms and measures of brain structure, in that presence of depression or anxiety or increased symptoms of the disorders were associated with diminished brain structure. However, some papers did find the opposite effects. For depression, both positive and negative relationships were found in the precuneus. Szymkowicz et al. (2017) examined cortical alterations of the precuneus in relation to symptom dimensions of depression. They found that a greater number of somatic symptoms were associated with greater cortical thickness in the right precuneus. They also found an age x somatic symptom interaction for the left precuneus where higher levels of somatic symptoms at younger ages were associated with increased cortical thickness and at older ages were associated with reduced cortical thickness. This demonstrates the importance of considering interacting variables such as age, which has a profound effect on brain structure, when examining these relationships.

They also found that depression symptom dimensions did not affect the relationship between age and surface area, suggesting that cortical thickness may be a more sensitive measure of brain abnormalities relating to depressive symptoms in middle-ages to older adults. They had previously found a positive relationship between depression symptoms and cortical thickness in the precuneus (Szymkowicz et al., 2016) and found that higher total depressive symptoms were associated with less age-related cortical thinning, driven by somatic symptoms. They could not explain the discrepancy between the two studies. Ries et al. (2009) also reported diminished brain structure in the precuneus in individuals with mild to moderate depressive symptoms compared with healthy controls. With regards to anxiety, as mentioned above, there seems to be a positive or negative relationship between trait anxiety and brain structure, specifically cortical thickness, in the presence or absence of a history of depression. Thus, at least in the anterior cingulate cortex, the insula and the orbitofrontal cortex, a history of depression seems to mediate the relationship between trait anxiety and cortical thickness and affect the direction of this relationship (Potvin et al.,2015).

We also aimed to establish whether these brain correlates were common across depression and anxiety or distinct. This review was not able to answer this question satisfactorily due to the small number of papers examining anxiety. There were five brain regions that were implicated in both anxiety and depression. These were the orbitofrontal cortex, the anterior cingulate cortex, the insula, the hippocampus, and the inferior frontal gyrus. Some of the associations found in the orbitofrontal cortex, the insula and the anterior cingulate cortex were contradictory across depression and anxiety. For example, in the orbitofrontal cortex, depression was associated with reduced GMV in two papers (Roussotte et al., 2017; Dotson et al., 2009) and reduced cortical thickness in two papers (Kwak et al., 2022; Lim et al., 2012) and anxiety was associated with both reduced and increased cortical thickness in another (Potvin et al., 2015). In the insula, reduced GMV and reduced cortical thickness were associated with increased depressive scores (Petkus et al., 2022; Tudorascu et al., 2014; Kwak et al., 2022) and increased and decreased cortical thickness was found for anxiety (Potvin et al., 2015). Finally, in the anterior cingulate cortex, increase in depressive scores has been associated with reduced GMV (Tudorascu et al., 2014), reduced cortical thickness (Kwak et al., 2022; Lim et al., 2012) and reduced FA (Tudorascu et al., 2014). Anxiety has also been associated with both reduced and increased cortical thickness, depending on the presence a history of depression (Potvin et al., 2015).

The majority of the papers in this study used ROI analyses rather than whole brain. On the one hand, one could argue that it is most informative and objective to use whole brain analysis as it is a data-driven approach and findings are not constrained by a priori expectations (Sexton et al., 2013). Also, it has been suggested that voxel-based morphometry (VBM), which is a whole brain analysis technique, allows tissue changes in areas that may be smaller and not defined as well anatomically to be identified more accurately (Ashburner & Friston, 2001). In addition, with ROI analysis, effects outside of these ROIs may go unnoticed (Sexton et al., 2013). On the other hand, VBM has come under criticism due to issues with the quality of its individual image outputs (Bookstein, 2001), and issues with quantifying group differences based on interactions between multiple brain networks (Gaonkar et al., 2011). It has been suggested that voxel-based statistics are decreased when group differences are “spatially complex and subtle” (Davatzikos, 2004). In addition, VBM requires correcting for multiple testing due to testing thousands of voxels (Gaonkar et al., 2011), and requires registering to standard template space, which can distort images. However, VBM has elucidated affected areas that may be missed by ROI. For example, Sexton et al. (2013) conducted a systematic review and meta-analysis and found differences in occipital and parietal regions using VBM, areas that are not often studied by ROI. This suggests that findings may be missed when guided by a priori hypotheses. There are, however, a number of reasons that ROI analysis might be chosen, three of which were detailed by Poldrack (2007). Firstly, one may simply want to explore one’s data, secondly, conducting ROI analyses can control for Type I error by limiting the number of statistical tests, and finally, one may wish to specifically test a region that has previously been functionally defined (Poldrack, 2007). Despite the benefits of ROI analysis, Poldrack (2007) warned against using results from these analyses to draw conclusions as they are biased by the ROI selection process (Poldrack, 2007). In the current review, there were several results that were found in both whole brain and ROI analyses. For depression, negative relationships were found with both whole brain and ROI analyses for areas including the orbitofrontal cortex, hippocampus, cingulate cortex, insula, and temporal gyrus. Positive relationships were found between the precuneus and depression in both ROI and whole brain analyses (Szymkowicz et al., 2017; 2016). The only studies that generated significant results when including anxiety in their brain analysis used ROI analysis. Arguably, given the benefits and problems with these two techniques, research should include both analyses, to ensure findings are not missed, but avoid applying both methods to the same dataset which could make the results vulnerable to confirmation bias.

There are several different mechanisms proposed in the papers within this systematic review to explain the relationships seen between depression and anxiety and changes in brain structure. Some of these proposed mechanisms may apply to depression or anxiety in general, or specifically to these disorders in later life. For example, Almeida et al. (2003) demonstrated that loss of frontal lobe volume and asymmetry of the frontal lobes was specific to late onset depression (LOD) and was not present among older adults with early onset depression (EOD). They suggest that their study supports the idea that disruption of fronto-subcortical circuitry is associated with the development of depression and that while the cause of reduced frontal lobe volume is not known, it cannot be simply attributed to natural ageing and could potentially be due to atrophy due to subcortical white matter changes or more subtle vascular changes in the presence of depression (Almeida et al., 2003). Sachs-Ericsson et al (2013) also compared LOD with EOD and found that participants with LOD had greater levels of cognitive decline than those with EOD and participants with EOD lost right hippocampal volume at a slower rate than those with LOD. However, when controlling for hippocampal volume changes over time, LOD was still found to predict a greater decrease in cognitive functioning than EOD. They suggest that this means that there must be additional neural mechanisms other than hippocampal atrophy that are affecting cognitive functioning in the LOD group. They were unable to identify the specific other neural mechanisms in this study but suggest that future research should investigate this. Zeki Al Hazzouri et al. (2018) also proposed a potential cerebrovascular mechanism for the relationship between depression and smaller cerebral volumes found in their study. They discuss the “vascular depression hypothesis” that suggests that cerebrovascular disease contributes to the development of depression, but that this relationship may be bidirectional (Zeki Al Hazzouri et al., 2018). In addition, they discuss neurodegenerative processes such as elevated cortisol and beta-amyloid plaque deposition as being potential candidates for the development of depression (Zeki Al Hazzouri et al., 2018). Another potential mechanism is proposed by Espinosa et al. (2022) for their finding that older adults with EOD had significantly larger bilateral thalamic volumes compared with a comparison group without a history of depression. They acknowledge that further work is needed to fully understand the mechanisms of these thalamic changes, however they suggest that persistent exposure to excitatory transmission from prefrontal cortical regions to thalamic nuclei, which has been found to show increased activity during depressive episodes, may lead to thalamic neurotoxicity (Espinosa et al., 2022). This neurotoxicity could in turn cause microdamage which may initiate a compensatory inflammatory response during remission of depressive symptoms, thus leading to the increased thalamic volumes that are seen (Espinosa et al., 2022). Thus, it could be suggested that there may be a number of different biological processes that may cause the same disorders. Therefore, identifying once specific brain biomarker for depression and anxiety may not be possible.

### Limitations of Studies in the Review

There were several limitations of the studies included in the final review that impacted on our ability to fully answer the research questions we set out prior to conducting the review. Firstly, most neuroimaging studies have relatively small sample sizes, and thus are not sufficiently powered to be able to fully elucidate the specific influence of other key clinical and demographic factors such as number of episodes, age at onset, medication effects, comorbidity, depression/anxiety sub-types etc. In this review specifically, many papers looked at subclinical depression/anxiety and thus information about age of onset, number of episodes and symptom duration is not necessarily even available. These variables have been shown to have an impact on brain measures and thus not having information on them, whilst understandable when subclinical populations are used, further adds to the heterogeneity of findings.

The review also highlighted the fact that research that investigates the relationship between depression and/or anxiety and brain structure in middle-aged to older adults does not acknowledge or investigate the role that social factors play in this relationship beyond simply using them as covariates. The majority of the papers (88.71%) measured at least one social factor, usually education, yet only eight papers discussed the effect that these factors may have on the variables of interest or the relationships between them. Given the evidence in the literature that social factors are strongly associated with depression and anxiety (e.g. McLaughlin et al., 2012; Chen et al., 2019; Motoc et al., 2019; Langlois et al., 2020; Leigh-Hunt et al., 2017; Álvarez et al., 2011; Chapman & Anda, 2007; McKenzie et al., 2013 and brain structure (e.g Walsh et al., 2014; McLaughlin et al., 2014; Jednoróg et al., 2012; Noble et al., 2015; Cavanagh et al., 2013), it would be beneficial to see future research that considers the impact of these factors fully. In addition, none of the papers reviewed performed their analyses separately by sex. Analysing separately by sex is necessary to enable us to establish whether the relationships between depression or anxiety and brain structure differ between males and females. For example, in their systematic review paper, Farhane-Medina et al., (2022) suggest that brain regions such as the hippocampus, amygdala, and prefrontal cortex that are relevant to emotions may hold the explanations for differences in anxiety between the sexes. It was suggested that they are dimorphic structures that behave differently in men and women. They also determined that the literature has mixed results with regards to the contribution of brain structures to sex differences in anxiety (Fahrane-Medina et al., 2022). Thus, these contradictory findings mean that it is important to examine this relationship separately by sex.

Given the variation in sample size, depression and anxiety measures, clinical and subclinical symptoms and the lack of control for correlated variables, it is unsurprising that the empirical data are inconsistent. In addition, the variability in imaging acquisition techniques and analysis pipelines makes direct comparisons challenging. One of our initial aims was so examine the extent to which research has examined both depression and anxiety together, given their comorbidity. Only a small minority of papers controlled for anxiety or depression when examining the other variable, which can be considered a limitation, given that these disorders, although often comorbid, are distinct. It would be impossible to draw strong conclusions about the impact of depression or anxiety separately on brain structure or the effect of brain structure on individuals’ depression or anxiety scores specifically, without controlling for the other variable. Support for this comes from the finding from Potvin et al. (2015) that when participants did not have a history of depression, higher trait anxiety was associated with larger cortical thickness in all their cortical ROIs, whereas participants with a depression history showed a relationship between higher trait anxiety and smaller cortical thickness in all ROIs. Given the fact that the presence of depression changed the relationship between anxiety and brain structure, it seems that consideration or controlling of the other disorder is vital. This could be an important direction for future literature. Future research should make use of the exciting asset of big data projects such as UK Biobank (2006), or the Human Connectome Project which will have sufficient power to detect relationships whilst controlling for confounding variables.

### Limitations of the Review Process

Although care was taken for the review to be robust and systematic, the search was limited to English Language sources and relevant sources in other languages may contain useful information that was excluded in this review. The review also did not include grey literature, thereby limiting the scope of coverage.

Overall, this systematic review has contributed to the literature on the relationship between depression and anxiety and brain structure in middle-aged to older adults, with some limits to the strength of the conclusions that can be drawn. One aim of this review was to establish the brain correlates of depression and anxiety in middle-aged and older adults. While there are some regions that have been implicated in multiple papers, mostly with a negative relationship suggesting reduced structural integrity in the presence of depression and anxiety, there is often still inconsistency between studies. As discussed, this is perhaps unsurprising given the variabilities described previously. The review highlights an urgent need for further research into the relationship between brain structure and anxiety in middle-aged and older adults given the discrepancy between the number of papers examining anxiety compared to depression, the fact that we have an ageing population and the prevalence of anxiety and impact it has on the wellbeing of individuals in this age group. The review also set out to determine whether structural brain correlates were common across depression and anxiety or distinct, however given the contradictory nature of the results across the papers, further research that examines both depression and anxiety and finds a way to capture the unique contributions that each of the disorders make will be needed to fully answer this question. Future research should utilise large-scale, longitudinal research projects with neuroimaging data, depression and anxiety measures, and detailed sociodemographic data to examine causal relationships between depression and anxiety and brain structure, whilst not ignoring the important influence of the many variables discussed in this review. Large imaging projects such as the UK Biobank, which plans to continue to add further neuroimaging data collection points in the future, will be a useful tool to provide insights into this area of research.

## Data Availability

All data produced in the present study are available upon reasonable request to the authors and are already contained in the papers included in the review, which are referenced clearly.

## Appendix

**Table A:** PRISMA checklist for the systematic review.

| Section and Topic | Item # | Checklist item | Location where item is reported |
| --- | --- | --- | --- |
| **TITLE** |  |  |  |
| Title | 1 | Identify the report as a systematic review. | Title |
| **ABSTRACT** |  |  |  |
| Abstract | 2 | See the PRISMA 2020 for Abstracts checklist. | Some items met-Abstract |
| **INTRODUCTION** |  |  |  |
| Rationale | 3 | Describe the rationale for the review in the context of existing knowledge. | Introduction |
| Objectives | 4 | Provide an explicit statement of the objective(s) or question(s) the review addresses. | Introduction |
| **METHODS** |  |  |  |
| Eligibility criteria | 5 | Specify the inclusion and exclusion criteria for the review and how studies were grouped for the syntheses. | Methods:<br>*Search strategy & selection criteria and Eligibility criteria, screening and article selection* |
| Information sources | 6 | Specify all databases, registers, websites, organisations, reference lists and other sources searched or consulted to identify studies. Specify the date when each source was last searched or consulted. | Methods:<br>*Search strategy & selection criteria* |
| Search strategy | 7 | Present the full search strategies for all databases, registers and websites, including any filters and limits used. | Methods:<br>*Search strategy & selection criteria* |
| Selection process | 8 | Specify the methods used to decide whether a study met the inclusion criteria of the review, including how many reviewers screened each record | Methods: |
|  |  | and each report retrieved, whether they worked independently, and if applicable, details of automation tools used in the process. | *Search strategy & selection criteria and Eligibility criteria, screening and article selection and Quality assessment* |
| Data collection process | 9 | Specify the methods used to collect data from reports, including how many reviewers collected data from each report, whether they worked independently, any processes for obtaining or confirming data from study investigators, and if applicable, details of automation tools used in the process. | Methods: *Search strategy & selection criteria and Eligibility criteria, screening and article selection and Quality assessment* |
| Data items | 10a | List and define all outcomes for which data were sought. Specify whether all results that were compatible with each outcome domain in each study were sought (e.g. for all measures, time points, analyses), and if not, the methods used to decide which results to collect. | Methods: *Search strategy & selection criteria and Eligibility criteria, screening and article selection* |
|  | 10b | List and define all other variables for which data were sought (e.g. participant and intervention characteristics, funding sources). Describe any assumptions made about any missing or unclear information. | N/A |
| Study risk of bias assessment | 11 | Specify the methods used to assess risk of bias in the included studies, including details of the tool(s) used, how many reviewers assessed each study and whether they worked independently, and if applicable, details of automation tools used in the process. | Methods: *Search strategy &* |
|  |  |  | *selection criteria and Quality assessment* |
| Effect measures | 12 | Specify for each outcome the effect measure(s) (e.g. risk ratio, mean difference) used in the synthesis or presentation of results. | N/A |
| Synthesis methods | 13a | Describe the processes used to decide which studies were eligible for each synthesis (e.g. tabulating the study intervention characteristics and comparing against the planned groups for each synthesis (item #5)). | Methods: Search strategy & selection criteria and Eligibility criteria, screening and article selection |
|  | 13b | Describe any methods required to prepare the data for presentation or synthesis, such as handling of missing summary statistics, or data conversions. | N/A |
|  | 13c | Describe any methods used to tabulate or visually display results of individual studies and syntheses. | Results: Study Selection |
|  | 13d | Describe any methods used to synthesize results and provide a rationale for the choice(s). If meta-analysis was performed, describe the model(s), method(s) to identify the presence and extent of statistical heterogeneity, and software package(s) used. | N/A |
|  | 13e | Describe any methods used to explore possible causes of heterogeneity among study results (e.g. subgroup analysis, meta-regression). | N/A |
|  | 13f | Describe any sensitivity analyses conducted to assess robustness of the synthesized results. | N/A |
| Reporting bias assessment | 14 | Describe any methods used to assess risk of bias due to missing results in a synthesis (arising from reporting biases). | N/A |
| Certainty assessment | 15 | Describe any methods used to assess certainty (or confidence) in the body of evidence for an outcome. | N/A |
| **RESULTS** |  |  |  |
| Study selection | 16a | Describe the results of the search and selection process, from the number of records identified in the search to the number of studies included in the review, ideally using a flow diagram. | Methods: Figure 1<br>Results: Study selection |
|  | 16b | Cite studies that might appear to meet the inclusion criteria, but which were excluded, and explain why they were excluded. | Results: *Study selection* |
| Study characteristics | 17 | Cite each included study and present its characteristics. | Results: *Table 1* |
| Risk of bias in studies | 18 | Present assessments of risk of bias for each included study. | N/A |
| Results of individual studies | 19 | For all outcomes, present, for each study: (a) summary statistics for each group (where appropriate) and (b) an effect estimate and its precision (e.g. confidence/credible interval), ideally using structured tables or plots. | Results: *Table 2* |
| Results of syntheses | 20a | For each synthesis, briefly summarise the characteristics and risk of bias among contributing studies. | N/A |
|  | 20b | Present results of all statistical syntheses conducted. If meta-analysis was done, present for each the summary estimate and its precision (e.g. confidence/credible interval) and measures of statistical heterogeneity. If comparing groups, describe the direction of the effect. | N/A |
|  | 20c | Present results of all investigations of possible causes of heterogeneity among study results. | N/A |
|  | 20d | Present results of all sensitivity analyses conducted to assess the robustness of the synthesized results. | N/A |
| Reporting biases | 21 | Present assessments of risk of bias due to missing results (arising from reporting biases) for each synthesis assessed. | N/A |
| Certainty of evidence | 22 | Present assessments of certainty (or confidence) in the body of evidence for each outcome assessed. | N/A |
| **DISCUSSION** |  |  |  |
| Discussion | 23a | Provide a general interpretation of the results in the context of other evidence. | Discussion |
|  | 23b | Discuss any limitations of the evidence included in the review. | Discussion |
|  | 23c | Discuss any limitations of the review processes used. | Discussion |
|  | 23d | Discuss implications of the results for practice, policy, and future research. | Discussion |
| **OTHER INFORMATION** |  |  |  |
| Registration and protocol | 24a | Provide registration information for the review, including register name and registration number, or state that the review was not registered. | N/A |
|  | 24b | Indicate where the review protocol can be accessed, or state that a protocol was not prepared. | N/A |
|  | 24c | Describe and explain any amendments to information provided at registration or in the protocol. | N/A |
| Support | 25 | Describe sources of financial or non-financial support for the review, and the role of the funders or sponsors in the review. | Yes |
| Competing interests | 26 | Declare any competing interests of review authors. | Yes |
| Availability of data, code and | 27 | Report which of the following are publicly available and where they can be found: template data collection forms; data extracted from included | N/A |
| other materials |  | studies; data used for all analyses; analytic code; any other materials used in the review. |  |

**Table B:**
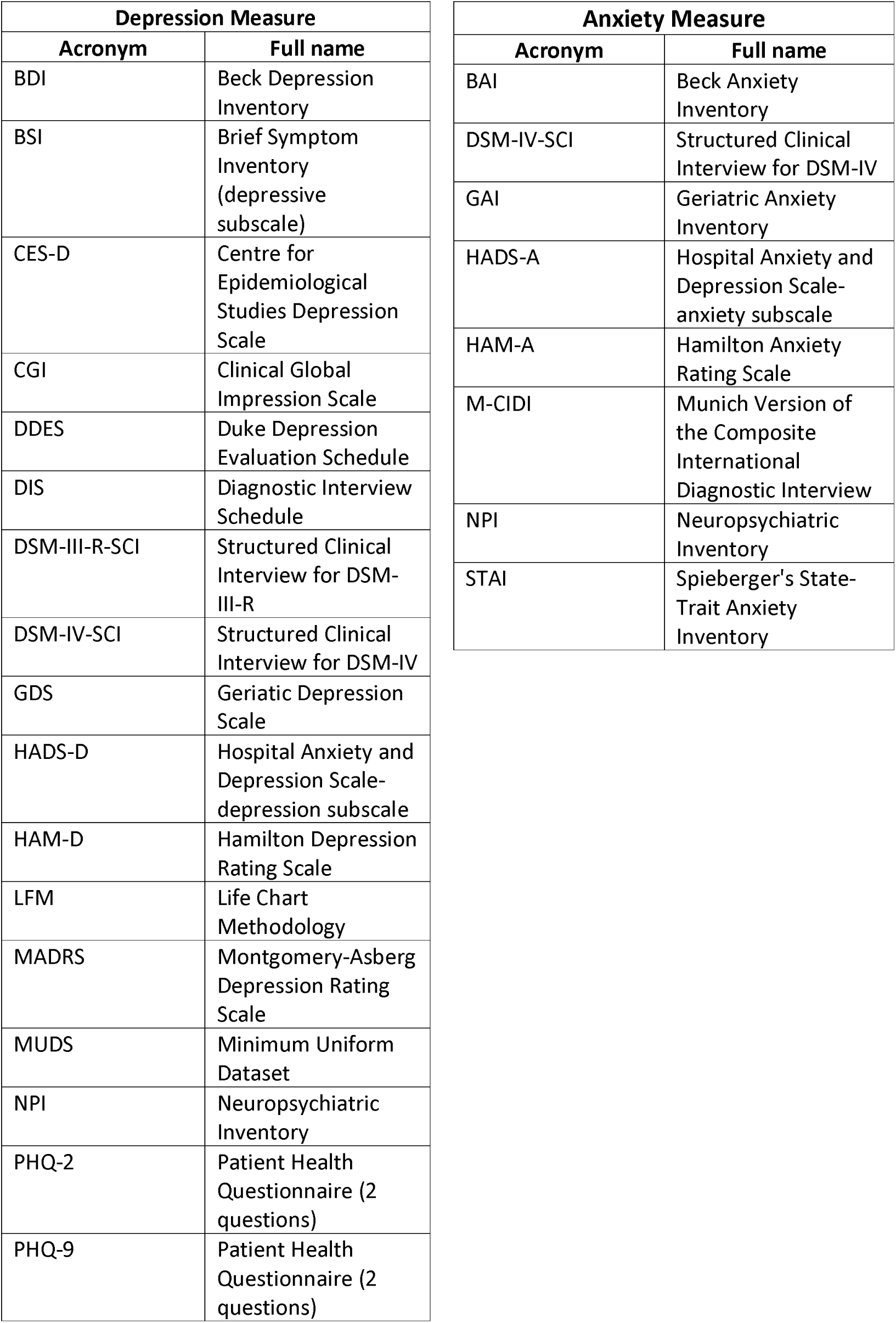
A glossary of depression or anxiety measures written as acronyms in the review paper.

